# Genomic epidemiology of SARS-CoV-2 transmission lineages in Ecuador

**DOI:** 10.1101/2021.03.31.21254685

**Authors:** Bernardo Gutierrez, Sully Márquez, Belén Prado-Vivar, Mónica Becerra-Wong, Juan José Guadalupe, Darlan da Silva Candido, Juan Carlos Fernandez-Cadena, Gabriel Morey-Leon, Rubén Armas-Gonzalez, Derly Madeleiny Andrade-Molina, Alfredo Bruno, Domenica de Mora, Maritza Olmedo, Denisse Portugal, Manuel Gonzalez, Alberto Orlando, Jan Felix Drexler, Andres Moreira-Soto, Anna-Lena Sander, Sebastian Brünink, Arne Kühne, Leandro Patiño, Andrés Carrazco-Montalvo, Orson Mestanza, Jeannete Zurita, Gabriela Sevillano, Louis du Plessis, John T. McCrone, Josefina Coloma, Gabriel Trueba, Verónica Barragán, Patricio Rojas-Silva, Michelle Grunauer, Moritz U.G. Kraemer, Nuno R. Faria, Marina Escalera-Zamudio, Oliver G. Pybus, Paúl Cárdenas

## Abstract

Characterisation of SARS-CoV-2 genetic diversity through space and time can reveal trends in virus importation and domestic circulation, and permit the exploration of questions regarding the early transmission dynamics. Here we present a detailed description of SARS-CoV-2 genomic epidemiology in Ecuador, one of the hardest hit countries during the early stages of the COVID-19 pandemic. We generate and analyse 160 whole genome sequences sampled from all provinces of Ecuador in 2020. Molecular clock and phylgeographic analysis of these sequences in the context of global SARS-CoV-2 diversity enable us to identify and characterise individual transmission lineages within Ecuador, explore their spatiotemporal distributions, and consider their introduction and domestic circulation. Our results reveal a pattern of multiple international importations across the country, with apparent differences between key provinces. Transmission lineages were mostly introduced before the implementation of non-pharmaceutical interventions (NPIs), with differential degrees of persistence and national dissemination.

## Introduction

The rapid generation of substantial numbers of virus genomic sequences during the COVID-19 pandemic is without precedent. During 2020, laboratories and institutes around the world produced and shared over 300000 whole SARS-CoV-2 genome sequences in the GISAID repository (1), providing an unparalleled data set that permits detailed analyses of virus transmission and dissemination. These achievements have provided insights into the sources of SARS-CoV-2 importation and early transmission dynamics in individual countries and geographical regions (2–5), and have enabled the exploration of viral transmission history at a global scale (6). Phylogenetic methods, including molecular clock models and phylogeographic and phylodynamic methods, are now used routinely to analyse such genomic data from emerging outbreaks (7). The resolution level of evolutionary and transmission history obtained using these methods is contingent on the virus’ evolutionary rate and the depth and representativeness of sampling of cases across space and time (8). While heterogeneous sampling and sequencing among countries can bias and affect the output of some phylogeographic methods (9, 10), general trends in the transmission of viral lineages can still be inferred from smaller samples of genomic sequences from individual locations.

The utility of pathogen genomic surveillance during outbreaks has developed during various past emerging epidemics (11–13) and has gained further momentum during the current global health crisis. Information about epidemiological trends can be effectively complemented with genomic analyses in order to understand case-specific (14) and general transmission patterns (15). This framework can be extended to account for other factors that affect the spread of pathogens, ranging from human mobility on a global scale (16) to particular social networks (17). The analysis of local-and national-scale data sets during the current pandemic has provided insights into the processes affecting the introduction and circulation of the virus into new locations (18) and provided genomic context for other data sources (19–21). Indeed, the integration and analysis of multiple data sources about an emerging epidemic has the potential to compensate for surveillance blind spots and better understand poorly sampled outbreaks (22).

The COVID-19 epidemic in Ecuador was marked by a dramatic and widely publicised early phase (23) with an estimated basic reproductive number (*R*_*0*_) of 3.54 (24). Ecuador is a small middle-income South American country with the seventh largest population in the continent; half of the country’s population lives in Guayas province (host of the country’s most populated city, Guayaquil) and Pichincha province (host of the country’s second most populated city, the capital Quito; Fig 1A). The first case was reported in the country on 27th February 2020 (a patient who returned from abroad through Guayaquil with date of symptom onset of February 15) and was followed by the declaration of a National Health Emergency on 11^th^ March 2020. Public health interventions were implemented shortly thereafter: mass gatherings were restricted on March 13, and a partial lockdown that included the closure of international borders was implemented on March 17. Finally, a full lockdown that included a curfew and the limitation of domestic mobility in private and public vehicles came into effect on March 25 (24). The country’s port city of Guayaquil was the first epicentre of the epidemic, facing a severe increase in the numbers of cases between late February and early April. The province of Guayas reached its highest effective reproductive number *R*_*t*_ (defined as the average number of secondary cases caused by a primary cases at a point in time *t*; 25) on March 14 (*R*_*t*_ estimates vary between 3.96 and 4.91) with 1462 cases reported that day (26) and reported a cumulative incidence of 146.94 cases per 100000 people by April 18 (24). The actual number of cases are likely to have been much higher when evaluated through the lens of excess mortality data (as obtained from the National Institute of Statistics and Census; Instituto Nacional de Estadística y Censos, INEC), and would explain why local diagnostic and healthcare services became rapidly overwhelmed (23, 24). After the peak and decline of the epidemic’s first wave in Ecuador, restrictions were maintained during April and May and progressively relaxed over the following months, as the epicentre of Ecuador’s epidemic moved to the capital city of Quito, located in Pichincha province (which on 23^rd^ July 2020 overtook Guayaquil as the city with the greatest number of COVID-19 confirmed cases). The last restrictions were finally lifted on September 13, although use of personal protective equipment and social distancing guidelines remained in place.

**Figure 1.**
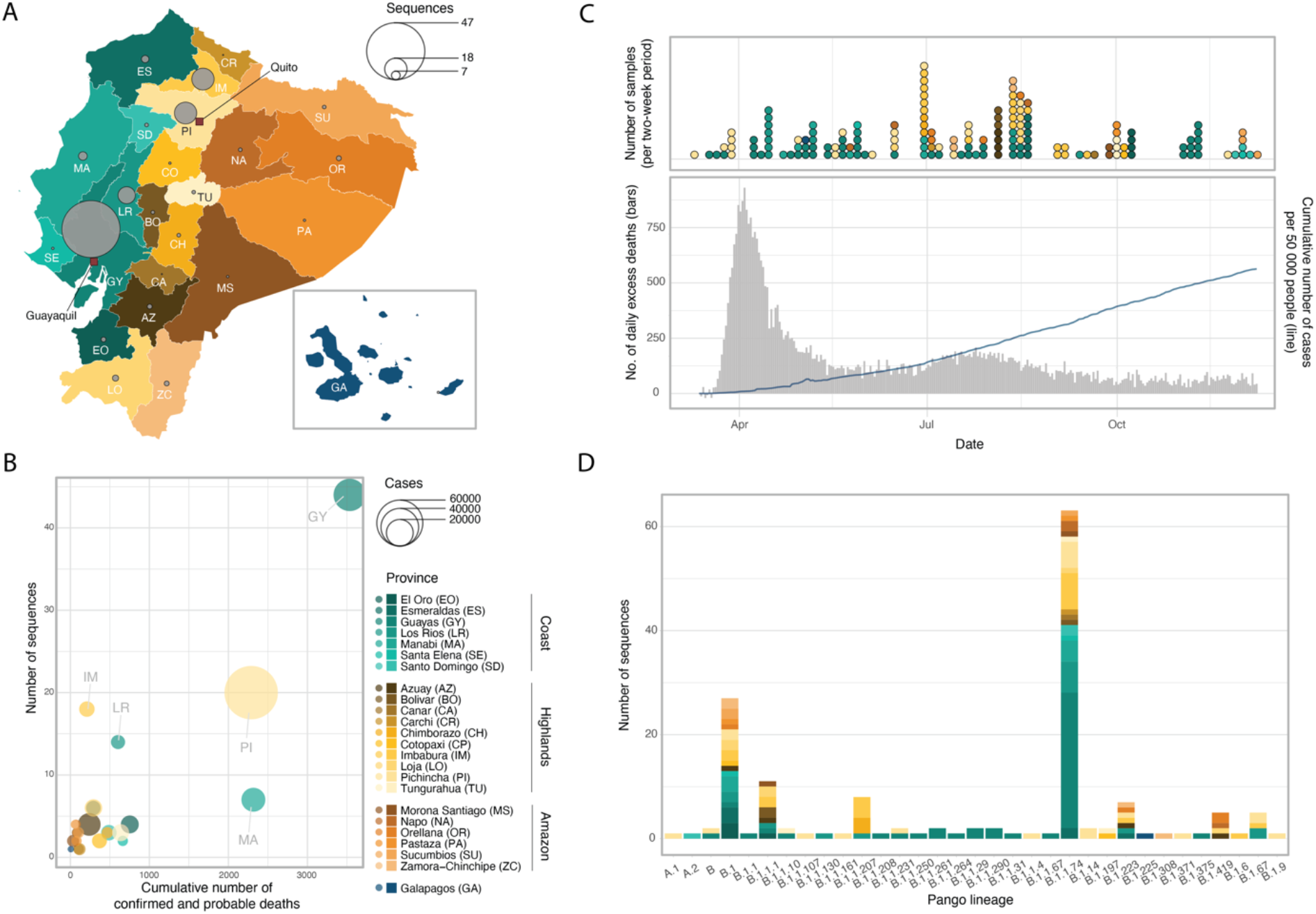
Overview of genomic sampling and SARS-CoV-2 genetic diversity in Ecuador. ***A***. Number of sequences from Ecuador analysed in this study per province across the four main geographic regions: the coast (shades of green), the highlands (shades of yellow), the Amazon (shades of orange) and the Galápagos (blue). ***B***. Number of deaths (attributed to laboratory-confirmed COVID-19 cases) versus number of whole genome sequences available per province. Circle radius shows the number of cases per province. ***C***. Timelines showing collection dates of sequences from the four geographic regions across time (upper panel) and the COVID-19 epidemiological curves in Ecuador during 2020 (cumulative number of laboratory-confirmed cases as reported by the Ministry of Health in the blue line, number of daily excess deaths compared to the same dates in 2019 as reported by the National Institute of Statistics and Census in grey; lower panel). ***D***. Geographic distribution of SARS-CoV-2 lineages identified in Ecuador.

To date, the source and diversity of circulating transmission lineages in Ecuador and their reach across the country remain unexplored. International importations are expected to have played an important role in seeding transmission chains in Ecuador, as observed in other countries (15). We undertake phylogenetic analyses of 160 SARS-CoV-2 whole genome sequences sampled from Ecuadorian cases and place them within the context of global viral genetic diversity in order to characterise the genomic epidemiology of SARS-CoV-2 in the country. We identify introduction events and transmission lineages within Ecuador and investigate their spatio-temporal distribution, and we hypothesise about the role of domestic seeding on viral transmission dynamics within Ecuador.

## Methods

### Genomic sequencing of SARS-CoV-2 samples from Ecuador

Clinical samples were collected from patients with a laboratory confirmed SARS-CoV-2 infection in Ecuador during 2020 (Fig 1A-C). Samples collected by the Microbiology Institute at Universidad San Francisco de Quito (IM-USFQ) were obtained from third-level hospitals (i.e. specialised tertiary referral hospitals) across all 24 provinces in the country without unified selection criteria (27). Samples collected by the Omics Sciences Laboratory at Universidad de Especialidades Espiritu Santo (UEES) were obtained from samples collected from the laboratory’s diagnostic service and selected at random for sequencing. Samples collected by the National Institute of Investigations in Public Health (Instituto Nacional de Investigación en Salud Pública, INSPI) were obtained from the national epidemiological SARS-CoV-2 surveillance system. Samples collected by the Biomedical Research Unit at Zurita & Zurita Laboratories (ZZL) were obtained from community patients from Quito who presented clinical signs of reinfection. This complete Ecuadorian sample set was collected between March 9 and December 9 2020, with limited representation during the early months of the epidemic when compared to excess mortality data (Fig 1C).

From these samples we generated 160 complete SARS-CoV-2 genomic sequences using different methodologies. IM-USFQ generated 108 whole genome sequences using Oxford Nanopore MinION sequencing and the ARTIC Network primer scheme approach as previously described (27). UEES generated 33 whole genome sequences through Illumina sequencing on a MiniSeq platform (Illumina, San Diego, CA). INSPI generated 15 sequences either in collaboration with Charité - Berlin University of Medicine (through Illumina sequencing) or on site at the Centre for Multidisciplinary Research of the Direction of Research, Development and Innovation (through Oxford Nanopore MinION sequencing as described in 28). Zurita & Zurita Laboratories generated 4 sequences through Illumina sequencing on a MiSeq platform (Illumina, San Diego, CA). Details of sample collection, sequencing and genome assembly are summarised in Table S1. Sample collection dates and the province-level geographical location of residence of the patient were included as metadata for all sequences in the country.

To determine the viral genetic diversity circulating in the country during the sampling period, all sequences from Ecuador were phylogenetically assigned under the global Pango lineage system using the *Pangolin v2.2.2* tool (https://virological.org/t/pangolin-web-application-release/482).

### Global SARS-CoV-2 data sets

The Ecuadorian virus sequences were analysed in the context of global SARS-CoV-2 genomic diversity by including all high-quality SARS-CoV-2 genome sequences and their accompanying metadata available in GISAID (1) on January 1 2021 (sequences were retained if they were > 29000 nucleotides long and < 5% of the sequence was missing). Sequences without a complete sample collection date or not attributed to human hosts were excluded, yielding a total of 218771 sequences from samples collected from December 1, 2019 up until December 10, 2020.

The large number of SARS-CoV-2 genomes generated during 2020 makes full-scale phylogenomic analyses computationally prohibitive. We therefore subsampled sequences from the abovementioned full data set (*i.e.* all GISAID sequences included in our analyses, excluding the complete set of Ecuadorian sequences) using two approaches. First, we randomly sampled one sequence per country per day from the full data set over the complete sampling period, to create a “*systematically-subsampled data set*” (comprised of 8606 sequences). In parallel, we arbitrarily generated three “*randomly-subsampled data sets*” consisting of 8606 randomly chosen sequences from the full data set, to match the size of the systematically-subsampled data set. These randomly-subsampled data sets were used to evaluate the performance of the background SARS-CoV-2 sequences as the genomic context for the identification of transmission lineages within Ecuador (see the *Phylogenetic identification of transmission lineages* section below). Finally, we added the sequences from Ecuador to each data set, resulting in a total of 8766 sequences per data set.

Each data set was aligned to the Wuhan-Hu-1 (GenBank accession: MN908947.3) reference genome sequence (29) using *Minimap 2.17* (30) to generate multiple sequence alignments. Sites containing >90% gaps relative to the sequences in their respective alignment were masked, whilst the untranscribed terminal regions (UTRs) were trimmed. After masking and trimming, the resulting alignments had a final length of 29,409 nucleotides, with the shortest partial genome sequences being cut down to 28,955 nucleotides long.

### Phylogenetic identification of transmission lineages

We followed a similar rationale and methodology to that described in du Plessis *et al* (15) to identify local transmission lineages. Phylogenetically linked sequences were inferred to have descended from a common ancestor if they were associated with a single inferred introduction event into Ecuador from an international location (5, 15). Ecuadorian transmission lineages therefore correspond to lineages of sequences sampled within the country that descend from a node inferred to have also occurred in Ecuador, which must in turn have descended from outside of the country. Given the unstructured sampling of the Ecuadorian sequences, some transmission lineages will likely correspond to epidemiologically linked cases (i.e. targeted investigation of epidemiological clusters); these have been identified as such in the text whenever the information was available.

Maximum likelihood (ML) phylogenetic trees were estimated from the systematically-subsampled data set and the randomly-subsampled data sets using *IQtree 2.1.1* (31) under a GTR+Γ substitution model. Node support was estimated through an SH-like approximate Likelihood Ratio Test (aLRT) (32). The tree for the systematically-subsampled data set was re-rooted by heuristically searching for the root placement that minimises the mean squared residual of a regression of sequence sample date against root-to-tip genetic distance, calculated using *TempEst v1.5.3* (33), to maximise the temporal signal of the data set. The same regression was used to assess the clock-like behaviour of the data set.

Subsequent analyses in our pipeline require an evolutionary rate estimation. We performed an exploratory analysis on a random selection of 866 genomes from the systematically-subsampled data set (~10% of the sequences, ensuring that representatives of the earliest and latest collection dates were included) in order to estimate the evolutionary rate of the data set over the sampling period. We used *BEAST v.1.10.4* (34) to obtain a clock rate estimate using the HKY substitution model and a strict molecular clock with a continuous-time Markov chain (CTMC) prior (35). We employed a Skygrid coalescent tree prior (36) that accounts for the 50 epidemiological weeks over which the genomes were sampled, plus a cut-off period that precedes the earliest collected SARS-CoV-2 sequences. Independent Markov Chain Monte Carlo (MCMC) chains were run for 40 million steps and subsequently combined after discarding the initial 10% of each run as a burn-in. Parameter convergence was assessed using the effective sample size (ESS) estimates of the combined chains using *Tracer v1.7.1* (37).

The systematically subsampled data set was analysed with *BEAST v1.10.5* (https://github.com/beast-dev/beast-mcmc/releases/tag/v1.10.5pre_thorney) using a newly implemented method that estimates the tree likelihood at each point of the MCMC chain. This approach takes a data tree (the aforementioned re-rooted ML tree) instead of an alignment and significantly reduces analysis time by using a simple model to rescale the ML phylogeny into a time-calibrated tree (see 38). Under this approach, the likelihood of each branch length is defined as a function of a Poisson distribution with a mean directly proportional to the clock rate (38, 39); we therefore used a rate of 6.28×10^−4^ substitutions/site/year, based on the median clock rate estimate obtained from our exploratory analysis. We also defined a coalescent Skygrid prior, similar to the one described for the exploratory analysis. A version of the data tree re-scaled to time by *BEAST* was used as a starting tree, and independent MCMC chains were run for 100 million steps and combined after discarding 10% of each run as burn-in.

To identify nodes associated with transmission lineages in Ecuador, we used a discrete phylogeographic model consisting of a two-state discrete trait analysis (DTA) (40) implemented in *BEAST v1.10.4* (34). Tips were assigned to one of two possible states (Ecuador vs non-Ecuador) and reconstruction of ancestral node states was undertaken using an asymmetric substitution model (40). The estimation of the source location for all nodes was performed on an empirical distribution of 500 time-calibrated trees obtained in the previous step. The expected number of DTA transitions between international locations and Ecuador were estimated using a robust counting approach (41). Two independent MCMC chains of 5 million steps each were combined for this analysis, after discarding the first 500000 steps of each run as burn-in. A Maximum Clade Credibility (MCC) tree was generated from the DTA posterior tree distribution by sampling 1000 trees from the combined MCMC runs in *TreeAnnotator* v.1.10. Each internal node was assigned a posterior probability for its inferred location, and these were used to evaluate uncertainty regarding the assignment of potential transmission lineages in Ecuador.

### Transmission lineages and transmission lineage groups

All phylogenetic clusters of sequences from Ecuador were inspected visually on the MCC tree to assign individual transmission lineages. The nomenclature of these country-specific transmission lineages followed a one-letter code in alphabetical order, defined by the earliest sample collection date in each transmission lineage. General features of each transmission lineage were summarised, such as the earliest and latest sample in each lineage, the number of provinces in which the transmission lineage had been identified and the number of sequences belonging to said lineage (used as a proxy of transmission lineage size). The consistency with which sequences were grouped into these transmission lineages was evaluated by visually inspecting the ML trees for the randomly-subsampled data sets and comparing the clusters of Ecuadorian sequences to those from the DTA analysis.

In two instances, transmission lineages within Ecuador clustered together in the DTA analysis into monophyletic groups that also included some international sequences (see *Results*). This topology includes the possibility that these groups of transmission lineages in fact correspond to single introduction events, misidentified by our analyses as multiple introductions. This might indeed be the case given the variation in intensity of SARS-CoV-2 sampling across countries (6, 9), including Ecuador, and the limited genetic divergence observed in SARS-CoV-2 over the time span being analysed (42). We therefore identified these pairs of transmission lineages under a single letter (highlighted with an asterisk) for summarisation purposes, and evaluated and discussed their individual trajectories further.

## Results

### SARS-CoV-2 genetic diversity in Ecuador

The samples from laboratory-confirmed individuals obtained across mainland Ecuador (and one sample from the Galápagos Islands) were collected as they became available through different hospitals and laboratories and yielded representative genomes from all provinces in the country (Fig 1A). The number of sequences per province correlates with the cumulative laboratory-confirmed COVID-19 cases (Spearman’s *rho* = 0.597, *p* = 0.002) and number of deaths of patients with a positive or suspected-positive COVID-19 PCR test over the sampling period (Spearman’s *rho* = 0.492, *p* = 0.015), suggesting that the number of sequences per province is proportional to the number of infections. Despite the limited number of sequences from Ecuador, the representativeness of our sample is similar to that of other countries in the region. We estimate Ecuador produced 8 sequences for every 10000 reported cases or 12 sequences for every 1000 officially reported COVID-19 deaths. This is better than Peru (4 sequences per 10000 cases/10 sequences per 1000 deaths) or Brazil (3 sequences per 10000 cases/10 sequences per 1000 deaths) but less representative than Uruguay (159 sequences per 10000 cases, and a higher number of SARS-CoV-2 genome sequences than reported deaths) (Fig S1, File S1). It should be noted however that these estimates rely on the testing intensities between provinces in Ecuador and between different countries; limited testing in Ecuador could mean that the overall representation is lower than estimable from official reports.

Wide variation in the total numbers of cases across provinces in Ecuador is reflected in variation in number of sequences obtained. While heavily affected provinces such as Pichincha (72,305 confirmed cases until December 10) and Guayas (26,080 confirmed cases) account for larger numbers of sequences (47 and 18 for Guayas and Pichincha respectively), less affected provinces in the southern Highlands (Azuay – 12,670 confirmed cases, and Loja – 7252) and the Amazon (Morona Santiago – 3422, Napo – 1605, Orellana – 2100, Pastaza – 2360 and Zamora Chinchipe – 1628) are represented by few sequences (28 in total). The single sequence obtained from the Galápagos corresponds to the low number of cases there. Manabí province appears to be underrepresented (14,061 confirmed cases) while Imbabura (5695 confirmed cases) and Los Ríos (4707 confirmed cases) are represented by higher numbers of genomes per death (Fig. 1B). Sequence sampling rates for each province (excluding Galápagos) varied between 3 and 86 sequences per 1000 deaths (File S2). The temporal distribution of samples collected in Ecuador during 2020 do not strongly match trends in reported excess deaths. More samples were collected in July and August, but fewer genomes were sampled in the early epidemic months (March to May) despite the high number of excess deaths reported then (Fig 1C). Sequence representation is greater for the coastal provinces during the early months of the epidemic (March to June), when the epicentre of the epidemic was based in the port city of Guayaquil (in the Guayas province, (23), and shifted towards higher sampling in the highlands and Amazon provinces, as the epicentre of the epidemic shifted towards the capital city of Quito (in Pichincha province) and as more cases were reported in the Amazon.

Virus genomes from Ecuador were assigned to specific Pango lineages (43) using the *pangolin* tool (https://virological.org/t/pangolin-web-application-release/482). The genomes were assigned to 33 different global lineages and predominantly B.1.1.74 (39.4% of all Ecuadorian sequences; Fig. 1D), one of the lineages descended from B.1.1 which became one of the most dominant lineages during the early phase of the pandemic in Europe and North America (after the virus was introduced from Asia; 3, 43). The geographic distribution of the SARS-CoV-2 lineage diversity in the country shows distinctive patterns. While the majority of the lineages observed at low-frequencies in Ecuador were found in Pichincha, the heavily affected (and highly populated) province of Guayas (where Guayaquil is located) exhibits a predominance of the B.1.1.74 lineage (59.1% of all sequences from this province; Fig 1D). B.1.1.74 is also abundant in the provinces of Los Ríos, which neighbours Guayas (49%), and Imbabura, which neighbours Pichincha (38.9%). Other common lineages, such as B.1 (16.9% of all sequences from Ecuador) and B.1.1.1 (6.9% of all sequences from Ecuador), are distributed across various geographical regions.

### Identification of Ecuadorian transmission lineages

We undertook exploratory phylogenetic analyses using different sequence subsampling schemes, as the exceptionally large number of available SARS-CoV-2 sequences prevents full analysis of the complete global data set. We estimated maximum likelihood (ML) trees of the Ecuador sequences in the context of different background data sets and performed Bayesian phylogenetic inference on a systematically-subsampled data set. The clustering patterns of Ecuadorian sequences in the ML trees showed some variation between data sets, but in the majority of cases remained consistent (File S3). We therefore derive our results from the systematically subsampled data set and discuss these in light of the randomly subsampled data sets.

We consistently found that a sizeable proportion of sequences from Ecuador did not cluster with other sequences from the country (54/160 sequences for the systematically subsampled data set, 48 to 51/160 for the randomly-subsampled data sets; File S3), and were therefore assigned as singletons and not associated with further virus spread within Ecuador detectable through genomic analysis. These singletons could in fact represent introduction events after which forward transmission in Ecuador did occur but this was not captured by the sample size of this study. We note that the majority of the singleton sequences were collected before mid-July (Fig S2); we speculate that they could represent predominantly early introduction events that occurred before the implementation of a national lockdown on March 16. While it is possible to establish a possible limit of dates on which each singleton was introduced, based on the last ancestral node inferred to have occurred outside of Ecuador, the precise importation date will fall somewhere between the inferred age of this preceding node and the collection date of the singleton sequence. The low sampling density in Ecuador and our subsampling schemes are likely to introduce uncertainty in estimating the age of these nodes and we therefore excluded these analyses from our results.

The remaining sequences (106/160 sequences for the systematically-subsampled data set) fell into two distinct categories. Firstly, 20 monophyletic clusters of Ecuadorian sequences were identified, capturing multiple introduction events and some local viral circulation patterns. These clusters were assigned to be separate Ecuadorian transmission lineages, named A through V (with exceptions detailed in the paragraph below). Each represents a single introduction event of the virus from an international destination, followed by local forward transmission in Ecuador (15).

Secondly, we identified two large monophyletic clusters that included sequences from international locations and Ecuador. These were not identified strictly as individual transmission lineages through our DTA approach, but rather as genetically similar groups of individual transmission lineages (Fig S3–S9). These results were likely driven by the phylogenetic placement of the non-Ecuadorian sequences resulting in the ancestral nodes being inferred to have existed outside of Ecuador (Fig 2A). While there is a possibility that these in fact represent multiple closely related yet independently introduced transmission lineages, we here label them as transmission lineage groups (highlighted with an asterisk, D* and H*) for summary purposes.

**Figure 2.**
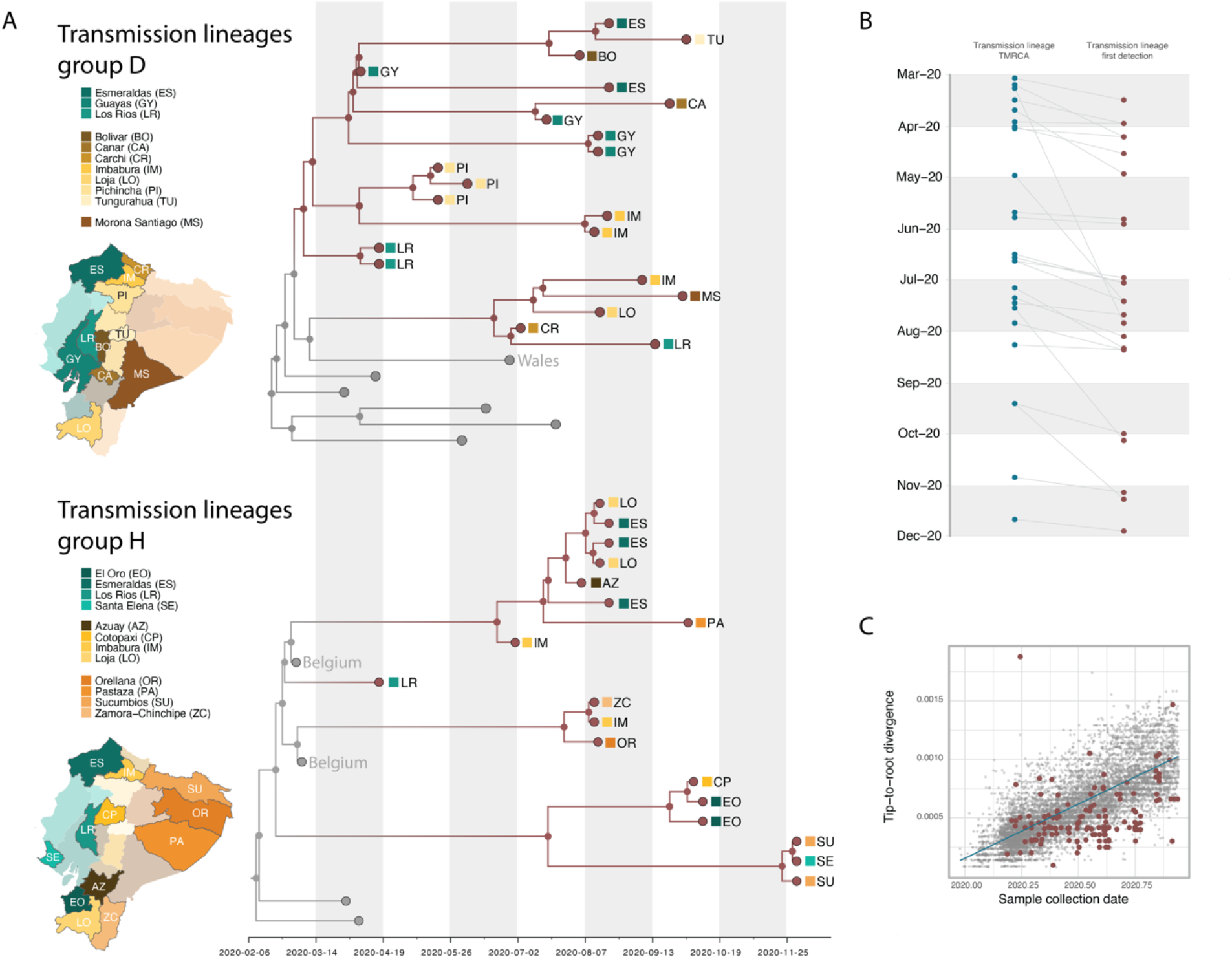
Time calibrated phylogenetic trees for the major transmission lineages in Ecuador. ***A***. Subtrees extracted from a time-calibrated Maximum Clade Credibility (MCC) tree of SARS-CoV-2 whole genome sequences, corresponding to the two largest clusters of sequences from Ecuador. Tree tips are coloured by sampling location (in Ecuador, red, versus outside of Ecuador, grey); nodes and branches are coloured by inferred location through a two-state DTA analysis. The province where each sequence was sampled is annotated on the tips, and maps highlight these provinces. Tips that correspond to sequences that cluster together within the major Ecuadorian clusters are also annotated with the region where the samples were collected. ***B***. Detection lag of individual transmission lineages in Ecuador, showing the median TMRCA of each transmission lineage from our data set (blue) connected by a grey line to the date of the earliest sequence in that transmission lineage (red). ***C***. Root-to-tip genetic distances (based on a heuristically rooted Maximum Likelihood tree) versus sample collection dates for the SARS-CoV-2 data set used in this analysis. Data points corresponding to sequences collected in Ecuador are highlighted in red, and the linear regression trendline is shown in blue.

Table 1 provides details for each Ecuador transmission lineage (named sequentially according to the collection date of the earliest sequence in each lineage). We identify 82 (95% HPD: 81-84) SARS-CoV-2 introduction events from other countries into Ecuador through a robust counting approach (41). This estimate assumes that transmission lineage groups D* and H* are comprised of 2 and 3 individual transmission lineages respectively (with an additional singleton inferred as part of H*; Fig 2A). The detection lag (defined as the number of days between the inferred transmission lineage TMRCA and its earliest sampled sequence) ranged between 1 and 140 days (Table 1), with a median of 16 days (IQR: 7-31 days; Fig 2B, Fig S10).

**Table 1.** Summary of transmission lineages identified in Ecuador.

| Transmission lineage | Number of sequences (% of total sequences) | Earliest sample collection date | Latest sample collection date | Median TMRCA (95% HPD) | Detection lag*** | Provinces where it has been sampled |
| --- | --- | --- | --- | --- | --- | --- |
| A | 3 (1.875%) | 16/03/2020 | 23/03/2020 | 03/03/2020<br>(25/02/2020 - 09/03/2020) | 13 days | Guayas |
| B | 5 (3.125%) | 30/03/2020 | 30/06/2020 | 16/03/2020<br>(02/03/2020 - 30/03/2020) | 14 days | Imbabura, Los Rios, Pichincha |
| C | 2 (1.25) | 30/03/2020 | 30/03/2020 | 29/03/2020<br>(26/03/2020 - 30/03/2020) | 1 days | Pichincha |
| D* | 21 (13.125%) | 07/04/2020 | 01/10/2020 | 02/03/2020**<br>(19/02/2020 - 14/03/2020) | 36 days** | Bolivar, Cañar, Carchi, Esmeraldas, Guayas, Imbabura, Loja, Los Rios, Morona Santiago, Pichincha, Tungurahua |
| E | 3 (1.875%) | 07/04/2020 | 30/06/2020 | 02/04/2020<br>(10/03/2020 - 07/04/2020) | 5 days | Guayas, Manabí |
| F | 6 (3.75%) | 13/04/2020 | 10/10/2020 | 12/07/2020<br>(02/06/2020 - 28/07/2020) | 23 days | Azuay, Bolivar, El Oro, Guayas, Loja |
| G | 8 (5%) | 17/04/2020 | 01/07/2020 | 01/04/2020<br>(25/02/2020 - 09/04/2020) | 16 days | Chimborazo, Imbabura, Los Rios |
| H* | 18 (11.25%) | 17/04/2020 | 30/11/2020 | 21/02/2020**<br>(11/02/2020 - 28/02/2020) | 56 days** | Azuay, Cotopaxi, El Oro, Esmeraldas, Imbabura, Loja, Orellana, Pastaza, Santa Elena, Sucumbios, Zamora Chinchipe |
| I | 3 (1.875%) | 29/04/2020 | 08/07/2020 | 22/03/2020<br>(24/02/2020 - 08/04/2020) | 38 days | Guayas |
| J | 2 (1.25%) | 26/05/2020 | 15/06/2020 | 22/05/2020<br>(03/05/2020 - 26/05/2020) | 4 days | Napo |
| K | 2 (1.25%) | 29/05/2020 | 20/07/2020 | 25/05/2020<br>(13/05/2020 - 29/05/2020) | 4 days | Guayas |
| L | 5 (3.125%) | 03/07/2020 | 19/08/2020 | 16/06/2020<br>(08/06/2020 - 02/07/2020) | 17 days | Azuay, Loja, Napo, Orellana |
| M | 3 (1.875%) | 14/07/2020 | 13/08/2020 | 30/04/2020<br>(26/03/2020 - 18/06/2020) | 75 days | Los Rios, Zamora Chinchipe |
| N | 7 (4.375%) | 14/07/2020 | 20/08/2020 | 20/06/2020<br>(19/05/2020 - 12/07/2020) | 24 days | Azuay, Esmeraldas, Imbabura, Los Rios, Orellana, Pichincha, Zamora Chinchipe |
| O | 3 (1.875%) | 22/07/2020 | 07/09/2020 | 15/07/2020<br>(02/07/2020 - 22/07/2020) | 7 days | Imbabura |
| P | 3 (1.875%) | 27/07/2020 | 04/11/2020 | 09/03/2020<br>(24/02/2020 - 08/04/2020) | 140 days | Guayas |
| Q | 2 (1.25%) | 11/08/2020 | 26/11/2020 | 06/07/2020<br>(22/05/2020 - 24/07/2020) | 36 days | Guayas, Pichincha |
| R | 2 (1.25%) | 12/08/2020 | 12/08/2020 | 09/08/2020<br>(20/07/2020 - 12/08/2020) | 3 days | Imbabura |
| S | 2 (1.25) | 01/10/2020 | 05/10/2020 | 13/09/2020<br>(16/08/2020 - 01/10/2020) | 18 days | Cotopaxi, Tungurahua |
| T | 2 (1.25%) | 05/11/2020 | 09/11/2020 | 27/10/2020<br>(05/10/2020 - 04/11/2020) | 9 days | Guayas |
| U | 2 (1.25%) | 09/11/2020 | 09/11/2020 | 13/09/2020<br>(16/08/2020 - 01/10/2020) | 57 days | Guayas |
| V | 3 (1.875%) | 28/11/2020 | 09/12/2020 | 21/11/2020<br>(01/11/2020 - 27/11/2020) | 7 days | Santo Domingo de los Tsachilas, Sucumbios |

\* Transmission complex, composed of two or more potential introductions of very genetically similar viruses.
\*\* TMRCA of various transmission lineages and sequences outside of Ecuador; node not inferred in Ecuador.
\*\*\* Number of days between the inferred TMRCA and the earliest collection date in the transmission lineage

### Size and persistence of transmission lineages

Initial molecular clock analyses showed our data set contained strong temporal signal overall, although many sequences from Ecuador showed lower than average genetic divergence from the root (Fig 2C). The inferred TMRCAs of Ecuadorian transmission lineages ranged from February to November 2020 (Table 1); from this list, transmission lineages C and S are composed of pairs of sequences that share an epidemiological link.

The TMRCAs estimated for the two large transmission lineage groups D* and H* are the earliest in our data, however, these might not represent true lineage ancestors within Ecuador, because each group could represent more than one introduction from other countries. After excluding these larger groups, we still identified six transmission lineages for which the 95% HPDs of the TMRCA include a time point that predates the implementation of the national lockdown, set on March 16. Therefore, these transmission lineages likely correspond to introduction events that occurred before restrictions on incoming international flights were adopted in Ecuador. An additional four transmission lineages have TMRCA estimates between late March and November 2020. These may correspond to more recent introductions (following the progressive relaxation of the lockdown in Ecuador between May and September). The most likely exceptions to this are transmission lineages C (TMRCA: 2020.2412, 95% HPD: 2020.2328-2020.2446) and M (TMRCA: 2020.3278, 95% HPD: 2020.1864-2020.4187). Incomplete sampling of these lineages and detection lags could result in the date of introduction being substantially earlier than the date of the TMRCA (44), which would place the introduction date for these transmission lineages prior to the implementation of the lockdown.

Transmission lineages varied in size from sequence pairs (transmission lineages C, J, K, Q, R, S, T and U) to larger clusters of 16 to 21 sequences (transmission lineage group D*, depending on whether D* is considered as a single lineage or as multiple lineages). The number of sequences in each transmission lineage was correlated with the number of days between the earliest and most recent sampling dates of sequences within the lineage (assuming that D* and H* are composed of multiple individual transmission lineages each). However, this result could be driven by the single largest transmission lineage in the data set. A similar pattern is observed when considering the time between the inferred TMRCA and the most recent sampled sequence of each transmission lineage (equivalent to the persistence time plus the detection lag; Fig S11). We also observed that lineages that were detected earlier tended to be larger (contain more sequences) and persisted longer but were not more geographically widespread (Fig 3A; Fig S12–S14). However, transmission lineages first detected between June and August did appear to be found in a greater number of provinces (Fig 3A). A similar pattern is observed when considering the TMRCA of each transmission lineage, where a greater persistence was also observed for lineages with a TMRCA between May and July (Fig 3A; Fig S13).

**Figure 3.**
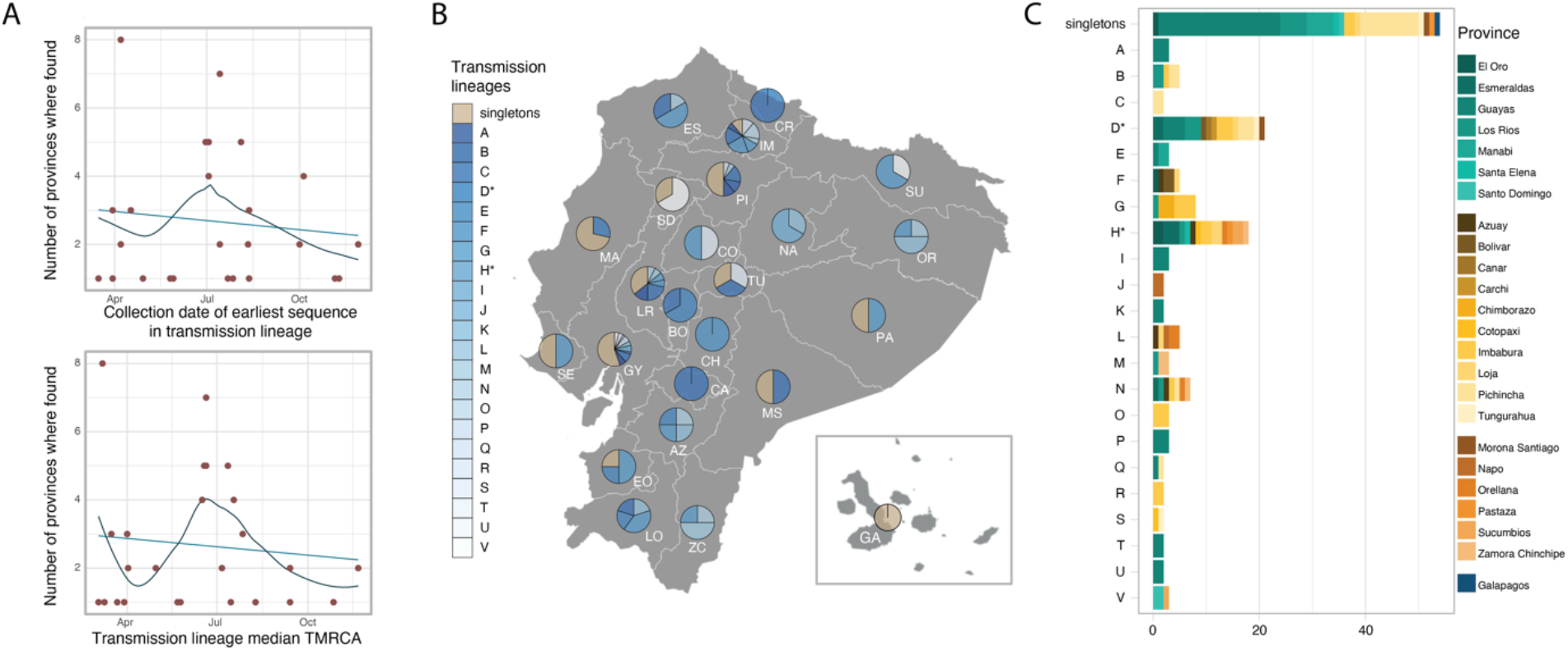
SARS-CoV-2 transmission lineages in Ecuador. ***A***. Summary of the geographic spread of transmission lineages in Ecuador, showing the number of provinces where each transmission lineage is found compared to the collection date of the earliest sequence in each transmission lineage (upper panel) or the inferred median TMRCA for each transmission lineage (lower panel). The trend lines show a linear regression in light blue and a fitted local polynomial regression in dark blue. ***B***. Contribution of individual transmission lineages and singleton sequences in each province. Transmission lineages (shades of blue) are ordered based on the earliest sample collection date in the group from earliest (darker) to more recent (lighter). ***C***. Bar plot summarising the provinces where each transmission lineage was sampled over the study sampling period.

### Geographical distribution of transmission lineages

Singletons and transmission lineages were found across multiple provinces and regions of Ecuador (Fig 3B-3C). Singletons represent an important proportion of the sequences in various provinces across central Ecuador ranging between 33.3% in Tungurahua and 52.3% in Guayas (Fig 3B), an observation that is particularly important for provinces with large numbers of sequences (Fig S14). On the other hand, different transmission lineages were found either in single provinces or across multiple regions (Fig 3C). The large transmission lineage groups D* and H* include sequences from provinces across three geographical regions each (the coastal region, the highlands and the Amazon region), and potentially show internal seeding events of the virus across provincial boundaries (Fig 2A, 3C). Even when accounting for the possibility that these lineage groups are comprised of multiple transmission lineages, sequences from different provinces and regions clustered together (Fig 2A).

Consistent with the spatio-temporal sampling patterns (Fig 1C), the older transmission lineages (shown in darker blue in Fig 3B) were identified predominantly in provinces on the coast and highland regions, while younger transmission lineages (shown in lighter blue in Fig 3B) have been identified in specific provinces in the north and more broadly in the south. The first epicentre of the COVID-19 epidemic in Ecuador, the province of Guayas, is represented by a high frequency of singleton lineages, with a high diversity of individual transmission lineages first identified at different times during 2020. A similar pattern is observed for the second epicentre of the epidemic, the province of Pichincha, but with fewer different transmission lineages and less representation of the youngest transmission lineages (Fig 3A). We note that these patterns could be affected by differences in the number of sequences available for each province (Fig 1A; Fig S15).

## Discussion

The early weeks of the COVID-19 epidemic in Ecuador were characterised by a severe spike in the number of cases in city of Guayaquil, the largest in the country located in the province of Guayas, and by high attack rates (*i.e.* new cases in a population at risk divided by the size of that population at risk) across various coastal provinces (24). The outbreak overwhelmed local healthcare systems, resulting in one of the highest excess death rates in the world during early 2020 (23). Information about the importation of SARS-CoV-2 into Ecuador and the domestic spread of the virus is needed to explain the drastic effects of the pandemic in the country during March and April 2020, and to explain the large difference in disease burden between Guayaquil and the capital, Quito.

An important determinant of the early dynamics of COVID-19 outbreaks has been human mobility and the number of introduction events of the virus into a new location with an immunologically naïve population, as was observed during the early stages of the emergence of SARS-CoV-2 in China (45). The large proportion of singletons observed in Guayas and various other coastal provinces, particularly given their tendency to occur early in the epidemic (Fig S2), could be suggestive of multiple independent introduction events with limited forward transmission. This would also explain why the earliest sequences in the local transmission lineages and the sequences assigned to the most common Pango lineage in Ecuador (B.1.1.74) were predominantly sampled in coastal provinces during the early weeks of the epidemic (Fig 2A, Fig S16).

Two additional factors support the hypothesis that Guayas played an important role in seeding of viral transmission to other regions in Ecuador: i) the city of Guayaquil hosts the second busiest international airport in the country and one of only two in the coastal region (the second international airport located in the province of Manabí hosts limited flights to a few international destinations; 46), and ii) the overall timing of the seeding events (which necessarily have to predate the inferred TMRCA of a lineage) corresponds to the school holiday period in the coastal region (February to April), when international travel and large social gatherings are more likely to occur. The inferred TMRCAs, which can serve as an upper bound for the true introduction dates of the largest lineages (assuming these are better represented with the available sequences), show that these transmission lineages most likely arrived in Ecuador before the date when NPIs were implemented and before travel restrictions came into place.

The genetic diversity of SARS-CoV-2 can be quantified using the Pango dynamic nomenclature system. Pango lineages reflect the history of significant events in the epidemic and geographic spread of the virus (47) and can be used to explore likely source locations of virus importations, for example, the high representation of lineages descended from B.1.1 in the province of Guayas (Fig 1D). The emergence of B.1.1 in Europe and North America around February 2020 (43) might suggest these regions contributed to seeding the epidemic in this province of Ecuador (despite the limitations on identifying exact source locations due to poor surveillance in many countries). B.1.1.74, the most prevalent lineage in Ecuador, is descended from B.1.1 and was more frequently sampled in Guayas during the early months of the epidemic (Fig S16), further revealing the role of locations where B.1.1 was prevalent as importation sources during the epidemic’s first wave. However, the high proportion of singletons observed in Guayas also suggests that onward transmission of introduced virus was less common. Insights from regions sampled at very high intensities, such as the United Kingdom, show that the majority of introductions lead to small, transient, dead-end transmission lineages, whereas a smaller number of introductions lead to larger and longer-lasting transmission lineages (15). If this phenomenon is a general property of the first wave of SARS-CoV-2 transmission, as appears to be the case given similar early observations in Brazil (5), Panama (48), Uruguay (49), Spain (50) and the Netherlands (51), we can expect that many of the introductions to Guayas led to few new cases, and that most of the ongoing transmission was derived from only a few introductions.

The larger transmission lineages identified here suggest that virus transmission was high between neighbouring and well-connected provinces. This might have been an important factor determining the transmission dynamics between the main cities in the country. In contrast to Guayaquil, Quito is the second-largest city in the country and presented a much less severe first epidemic wave, despite hosting the busiest international airport in the country. The city is located in the province of Pichincha, which exhibits a large proportion of singletons but fewer distinct transmission lineages overall. Pichincha also shows a lower diversity of global lineages and fewer different transmission lineages; furthermore, the transmission lineages observed in Pichincha are mostly not shared with Guayas. This suggests that either independent international introductions or domestic seeding events likely drove the early epidemic in Pichincha. Our phylogenetic analyses suggest that some transmission lineages in Pichincha were introduced from other provinces (Fig 2A, e.g. the monophyletic Pichincha clade in lineage group D*; Fig S6), suggesting that domestic travel might have played an important role in the establishment of SARS-CoV-2 transmission in this region. While it is also possible that a later burst of introductions of new lineages occurred from locations abroad, once international travel and lockdown measures were lifted, the sampling dates of singleton genomes from Pichincha (between March and July) suggest that introductions into Pichincha likely occurred before the lockdown rather than during the relaxation of NPIs. The Pichincha singletons account for the earliest sequences of this kind in our data set, but could represent instances where limited or no additional spread occurred following their introduction.

Ultimately, more comprehensive analyses on the sources and drivers of transmission would require a deeper sampling of key locations where transmission was high, and the inclusion of complementary data sources such as real-time mobility and transportation data, could provide a better overview of the forces shaping the observed viral genetic diversity in Ecuador. Provinces such as Azuay, Guayas and Pichincha represent the main air travel entry points, but the role of land mobility across the northern border with Colombia and the southern border with Peru should also be considered to further understand the role of other Latin American countries in regional viral transmission.

Our analysis highlights some important patterns but is limited by various factors. Most notably, the number of genome sequences in our study, although large by historical standards, is small compared to the new expectations set for virus genome sequencing during the COVID-19 pandemic. The sample size restricts our ability to infer further details about local virus transmission dynamics from sequence data alone. The trajectories of individual lineages, from their international sources to their spread across the country and their subsequent local circulation, are best analysed from larger data sets, or in conjunction with additional data sources to manage and ameliorate the potential consequences of sampling biases (10, 52). Incorporating data from self-reported travel histories and human mobility can help to maximise the utility of smaller samples, collected in settings where the sequencing of large numbers of genomes lies beyond local technological or financial capacity, or where high sampling densities are unfeasible (e.g. in remote locations).

The first year of the COVID-19 pandemic has shown how global connectivity plays a key role in the development of national epidemics caused by respiratory viruses, reminiscent of other pathogens such as Influenza viruses (16). Our results from Ecuador showcase the relevance of importations in establishing local viral circulation and the potential consequences of interprovincial mobility for highly connected locations. In particular, it shows that importations have been a common occurrence even after the implementation of lockdown measures and travel restrictions, and that seeding events across provinces can occur frequently. While air travel is limited between provinces, the connectivity provided by land travel can serve as a means for pathogen spread, highlighting the vulnerability of highly connected and remote locations alike. The notion that two large cities with busy international airports can manifest such different transmission dynamics and viral genetic diversity is also relevant, as it shows that a combination of multiple factors determines the outcome of an epidemic within a specific location. Ultimately, the type of interventions chosen to mitigate this high degree of connectivity, the necessity of an early implementation of these interventions and the adherence to these by the general population are paramount in determining their efficacy.

## Data Availability

The newly generated sequences have been publicly shared through the GISAID platform. The data sets (including Ministry of Health and National Institute of Statistics and Census data) and code used to generate the analyses presented in this study, as well as a list of accession numbers for the Ecuador SARS-CoV-2 genome sequences analysed here are
available through GitHub at https://github.com/BernardoGG/SARS-CoV-2_Genomic_lineages_Ecuador. We thank Andres N. Robalino and Carlos Oporto for their contribution to the open licence Ecuacovid repository (https://github.com/andrab/ecuacovid) used to extract epidemiological data for this study.

## Acknowledgements

We would like to thank all the clinical personnel from numerous public and private healthcare institutions who provided access to laboratory-confirmed samples to generate SARS-CoV-2 genomic sequences. We also thank the Centre for Arbovirus Discovery, Diagnosis, Genomics and Epidemiology (CADDE), Eva Harris from the A2CARES network and Lourdes Torres, Diego Quiroga and Stella de la Torre for their methodological, logistical and financial support (through the USFQ Emergency Grants) towards the sequencing of viral genomes. We are grateful to all laboratories and institutions worldwide involved in the generation of virus genome data shared on GISAID. Financial support was provided by the Clarendon Fund and the Department of Zoology of the University of Oxford (D.S.C.); WT fellowship 204311/Z/16/Z and MRC-FAPESP awards MR/S0195/1 and 18/14389-0 (N.R.F.); Branco Weiss Fellowship and EU grant 874850 MOOD (M.U.G.K.); WT Collaborators Award 206298/Z/17/Z (J.T.M.); Leverhulme Trust ECR Fellowship ECF-2019-542 (M.E.Z.); the Oxford Martin School (O.G.P., M.U.G.K. and L.d.P.); and the NIH Global Health Equity Scholars award FIC D43TW010540 (P.C.). The authors report no conflicts of interest.

## Data availability

The newly generated sequences have been publicly shared through the GISAID platform. The data sets (including Ministry of Health and National Institute of Statistics and Census data) and code used to generate the analyses presented in this study, as well as a list of accession numbers for the Ecuador SARS-CoV-2 genome sequences analysed here are available through GitHub at https://github.com/BernardoGG/SARS-CoV-2_Genomic_lineages_Ecuador. We thank Andrés N. Robalino and Carlos Oporto for their contribution to the open licence Ecuacovid repository (https://github.com/andrab/ecuacovid) used to extract epidemiological data for this study.

## Supplementary Information

**Table S1.** Summary of sample collection and sequencing methods used by different laboratories in Ecuador.

| Laboratory | Sample collection | Whole genome sequencing | Genome assembly |
| --- | --- | --- | --- |
| Institute of Virology, Charité-Universitätsmedizin Berlin | Nasopharyngeal swab samples were collected by the SARS-CoV-2 epidemiological surveillance system which were received at the National Reference Center for Influenza and other Respiratory Viruses at INSPI, under cold chain conditions. | Whole-genome amplification was done using the Artic Consortium PCR-based protocol (https://artic.network/ncov-2019). Library preparation and Illumina MiSeq sequencing were done using the KAPA Frag kit and KAPA Hyper Prep kit (Roche Molecular Diagnostics, Switzerland) and MiSeq reagent v2 chemistry (Illumina, USA) according to the manufacturers' protocols. | A modified pipeline dark-matter v3.1.88 (https://pypi.org/project/dark-matter/), using the tools AdapterRemoval, Bowtie2, samtools, gatk4, bcftools, mafft and ivar was used. In addition, the *de novo* assembly produced by Geneious software to the reference SARS-CoV-2 genome Wuhan-Hu-1 (GenBank accession number NC_045512.2) using default, was used to cross-validate as an internal control. |
| Centre for Multidisciplinary Research of the Direction of Research, Development and Innovation, National Institute of Investigations in Public Health (INSPI) | Nasopharyngeal swab samples were collected by the SARS-CoV-2 epidemiological surveillance system which were received at the National Reference Center for Influenza and other Respiratory Viruses at INSPI, under cold chain conditions. | See Lopez-Alvarez *et al.*, 2020. | See Lopez-Alvarez *et al.*, 2020. |
| Omics Sciences Laboratory, Faculty of Medical Sciences, Universidad de Especialidades Espíritu Santo (UEES) | Nasopharyngeal swabs preserved in 400 µL of 1X DNA/RNA Shield™ solution (Zymo Research, USA); saliva, urine, blood and semen samples were used for RNA extraction by Quick RNA viral kit (Zymo Research, CA, USA) according to the manufacturer's protocol in a biosafety type II chamber with HEPA filters. The RNA extracted was screened for the presence of SARS-CoV-2 genetic material by reverse transcriptase quantitative PCR (RT-qPCR) using Allplex™ SARS-CoV-2 Assay, a multiplex real-time PCR assay to detect 4 target genes: E, N, RdRp and S (Seegene Inc, CA, USA). | Nasopharyngeal swab RNA samples were further analyzed with the CleanPlex® SARS-CoV-2 FLEX Panel (Paragon Genomics, Hayward, CA, USA) following the manufacturer's protocol and sequenced on a MiniSeq platform (Illumina, San Diego, CA, USA) with a depth of 1000X. | Assembly and annotation procedures were performed in SOPHIA-DDM-v4. |
| Instituto de Microbiología, Universidad San Francisco de Quito (IM-USFQ) | Bronchi-alveolar lavage (BAL), nasopharyngeal swabs; see Márquez *et al.*, 2020. | See Márquez *et al.*, 2020. | See Márquez *et al.*, 2020. |
| Biomedical Research Unit, Zurita & Zurita Laboratorios (ZZL) | Nasopharyngeal swab samples from patients with suspected re-infections. | Whole genome sequencing of the SARS-CoV-2 samples was performed using the Paragon Genomics CleanPlex® SARS-CoV-2 Panel following the manufacturer's instructions on a MiSeq platform (Illumina, San Diego, CA, USA). | FastQC was used to assess the raw reads' quality (Sah *et al.*, 2020) before cleaning them with Trimmomatic 0.39 (PE - phred33 ILLUMINACLIP: TruSeq3-PE.fa:2:30:10:2:keepBothReads (Tillett *et al.*, 2020), LEADING:20 TRAILING:20 SLIDINGWINDOW:4:20 MINLEN:40) (Sah *et al.*, 2020). Clean reads were mapped to the reference SARS-CoV-2 genome Wuhan-Hu-1 (GenBank accession number NC_045512.2) (Sah *et al.*, 2020; Tillett *et al.*, 2020), using BWA-MEM 0.7.17 with default parameters (Sah *et al.*, 2020). PCR duplicates were marked and removed with Picard 2.23.8 (Tillett *et al.*, 2020). Variant calling was performed with bcftools 1.11 (-q 25 -Q 35) (Li *et al.*, 2020). Only high-quality variants (QUAL ≥ 20 and DP ≥ 5) (Karamitros *et al.*, 2020; Tillett *et al.*, 2020), were retained and used to generate consensus genomic sequences. In addition, the *de novo* assembly produced by MEGAHIT 1.2.9 using default parameters (Sah *et al.*, 2020), was used to cross-validate with the reference-based method as an internal control using blastn (Johnson *et al.*, 2008). |

---

**Fig S1.**
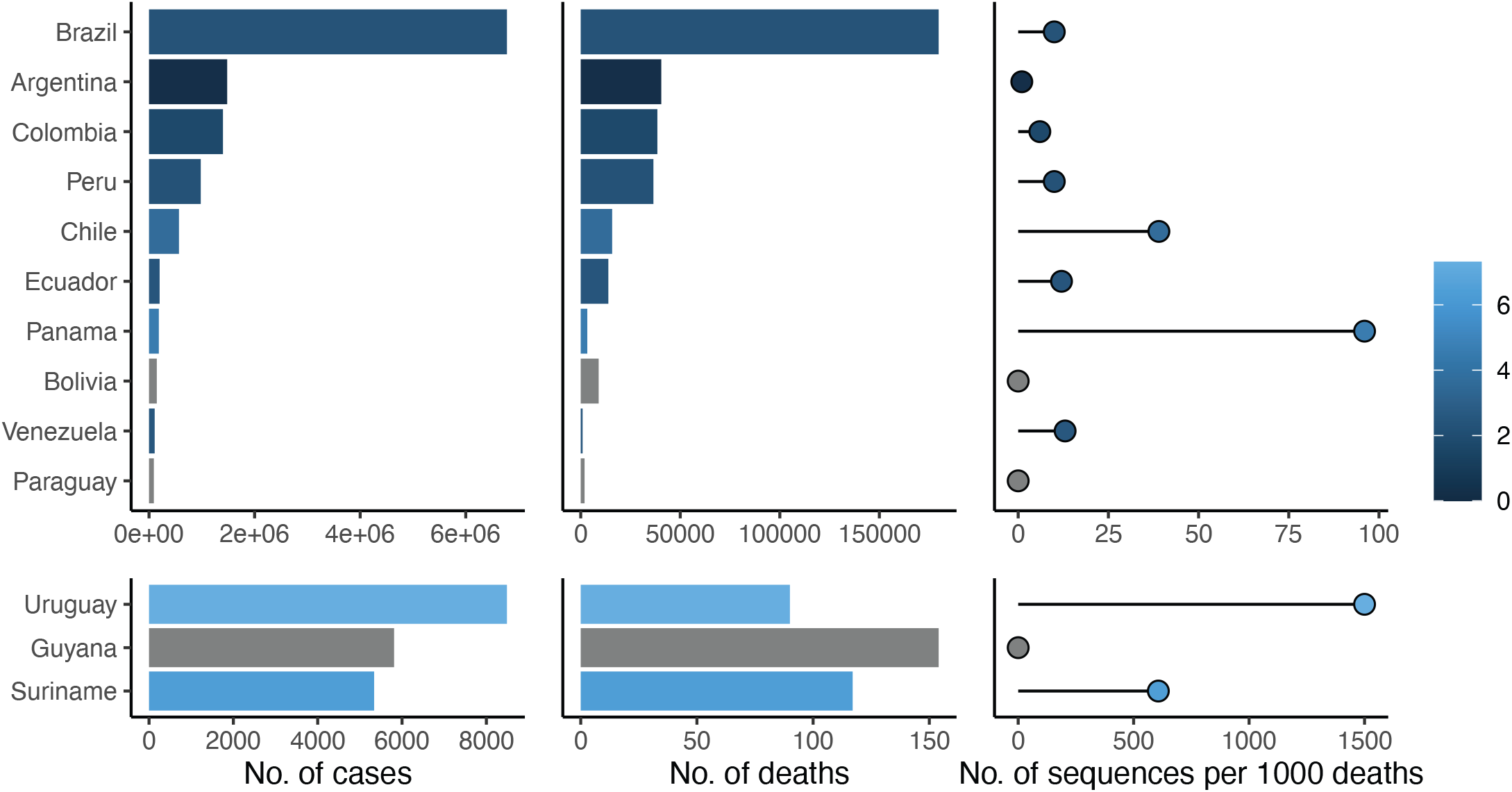
Comparison of the reported numbers of cases and deaths and the publicly available SARS-CoV-2 genomes generated per country in South America. Numbers of sequences were taken from the GISAID database, and the reported numbers of cases and deaths were taken from Our World in Data. Data retrieved on February 14 2021, with a cutoff date on 2020-12-10. Lower panels show data from countries at a different scale compared to the upper panels for visualisation purposes. Color scheme shows the (logarithmic) numbers of sequences per cumulative number of reported death.

**Fig S2.**
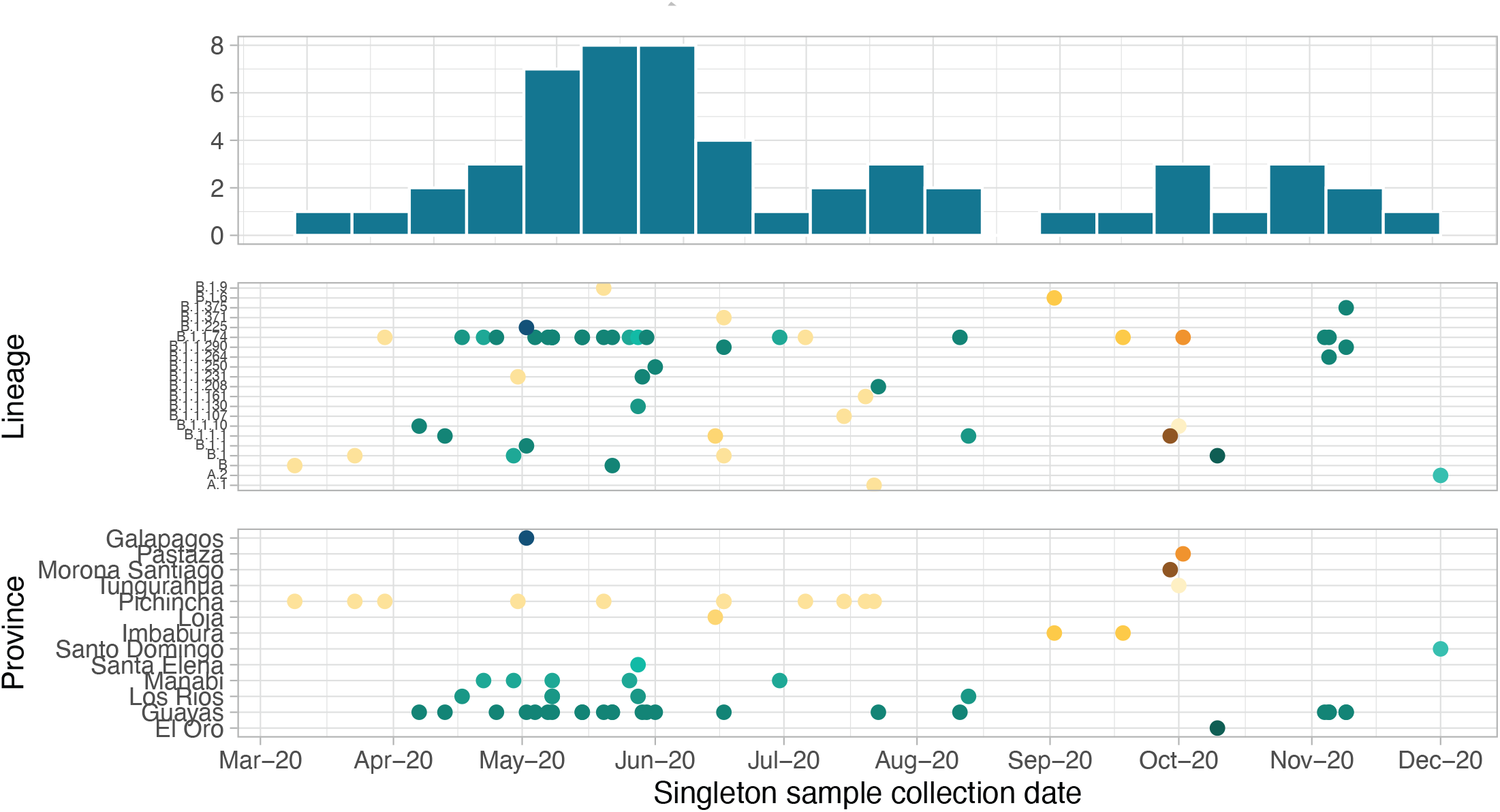
Summary of singletons identified in Ecuador. The upper panel shows the number of singletons per two-week epidemiological weeks. The middle panel shows the Pango lineages to which singletons were assigned, and the lower panel shows the province where singletons were identified by sample collection date. Dot colours correspond to the province where the samples were collected.

**Fig S3.**
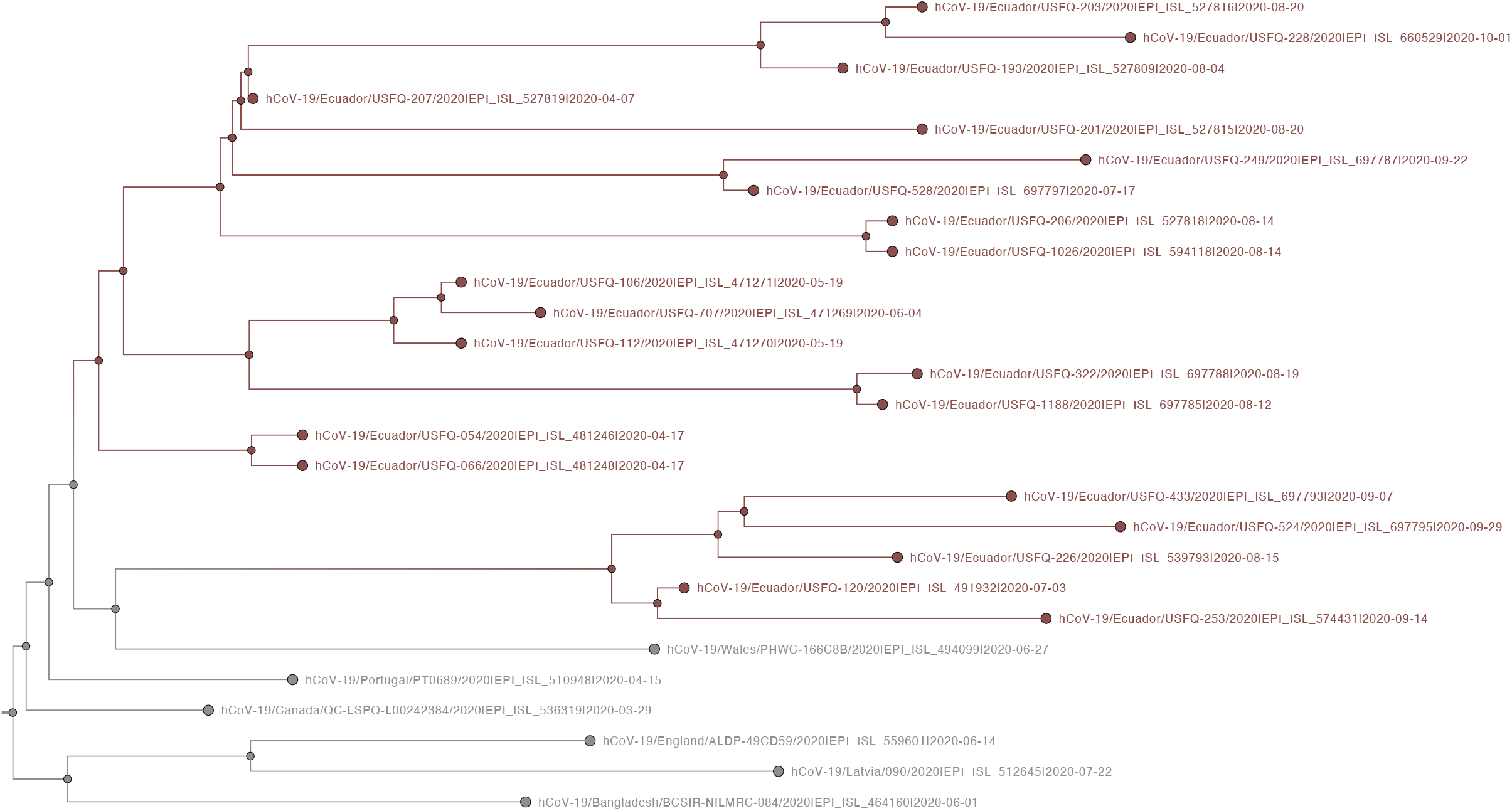
Subtree from the MCC tree, corresponding to transmission lineages group D.

**Fig S4.**
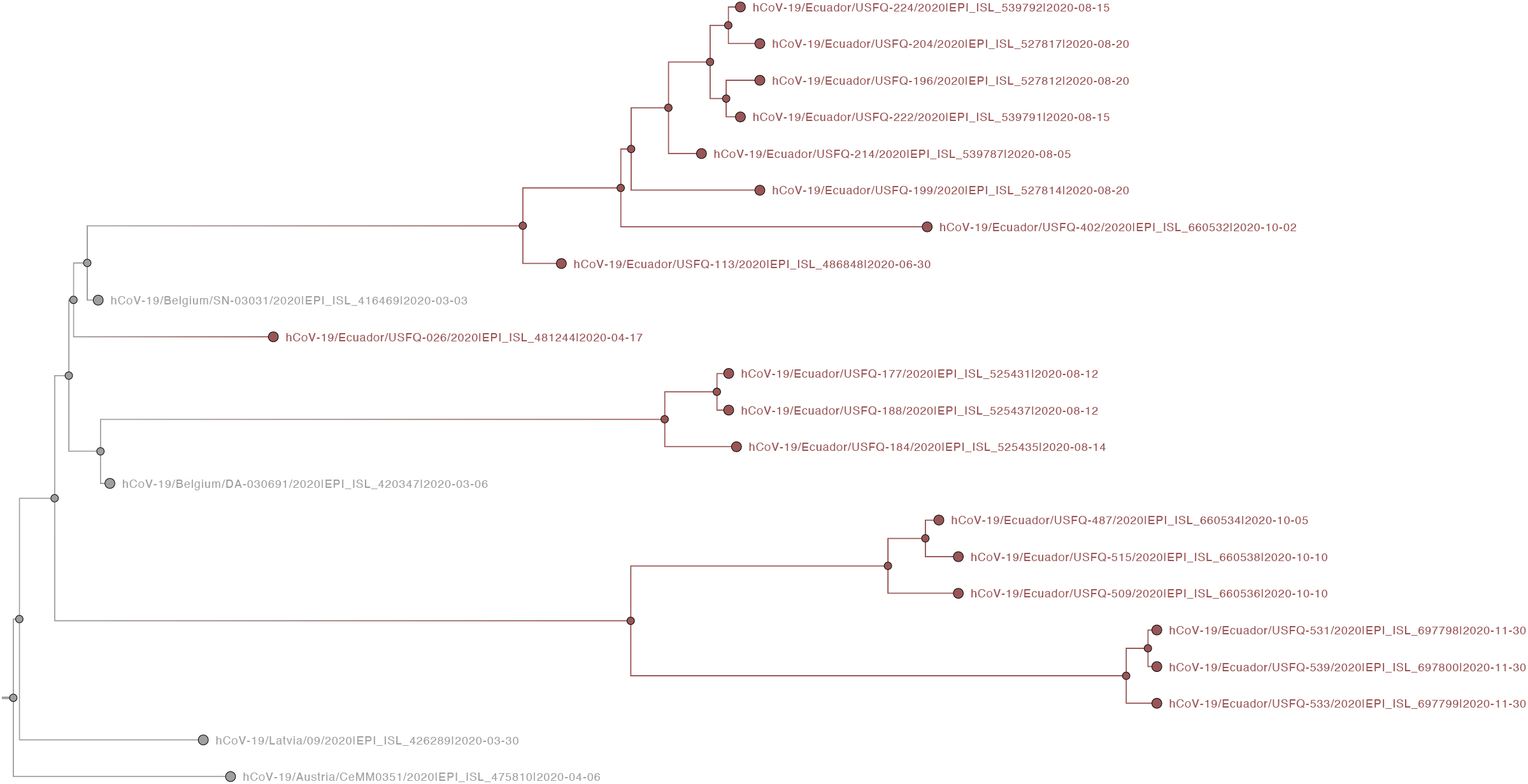
Subtree from the MCC tree, corresponding to transmission lineages group H.

**Fig S5.**
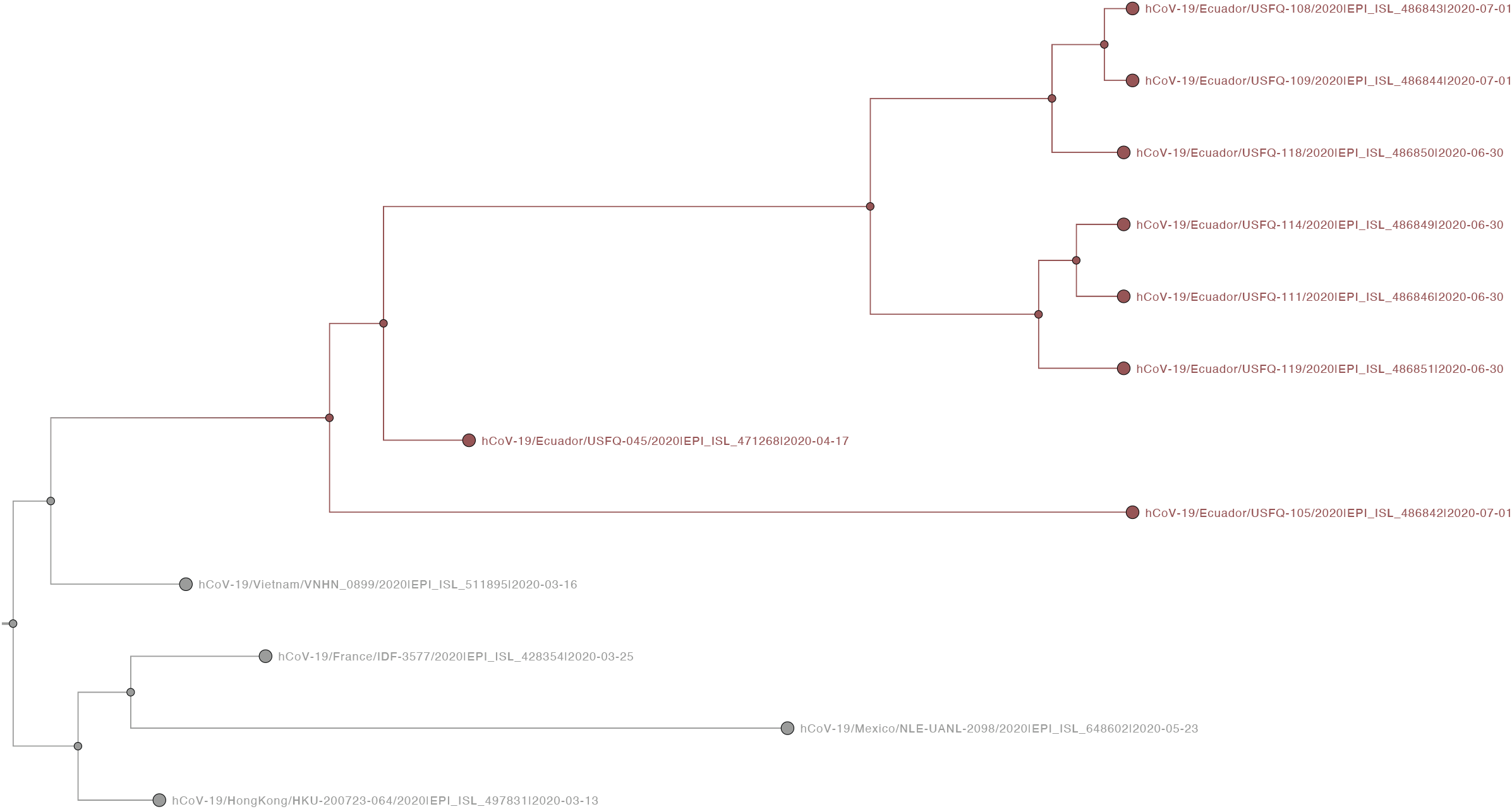
Subtree from the MCC tree, corresponding to transmission lineage G.

**Fig S6.**
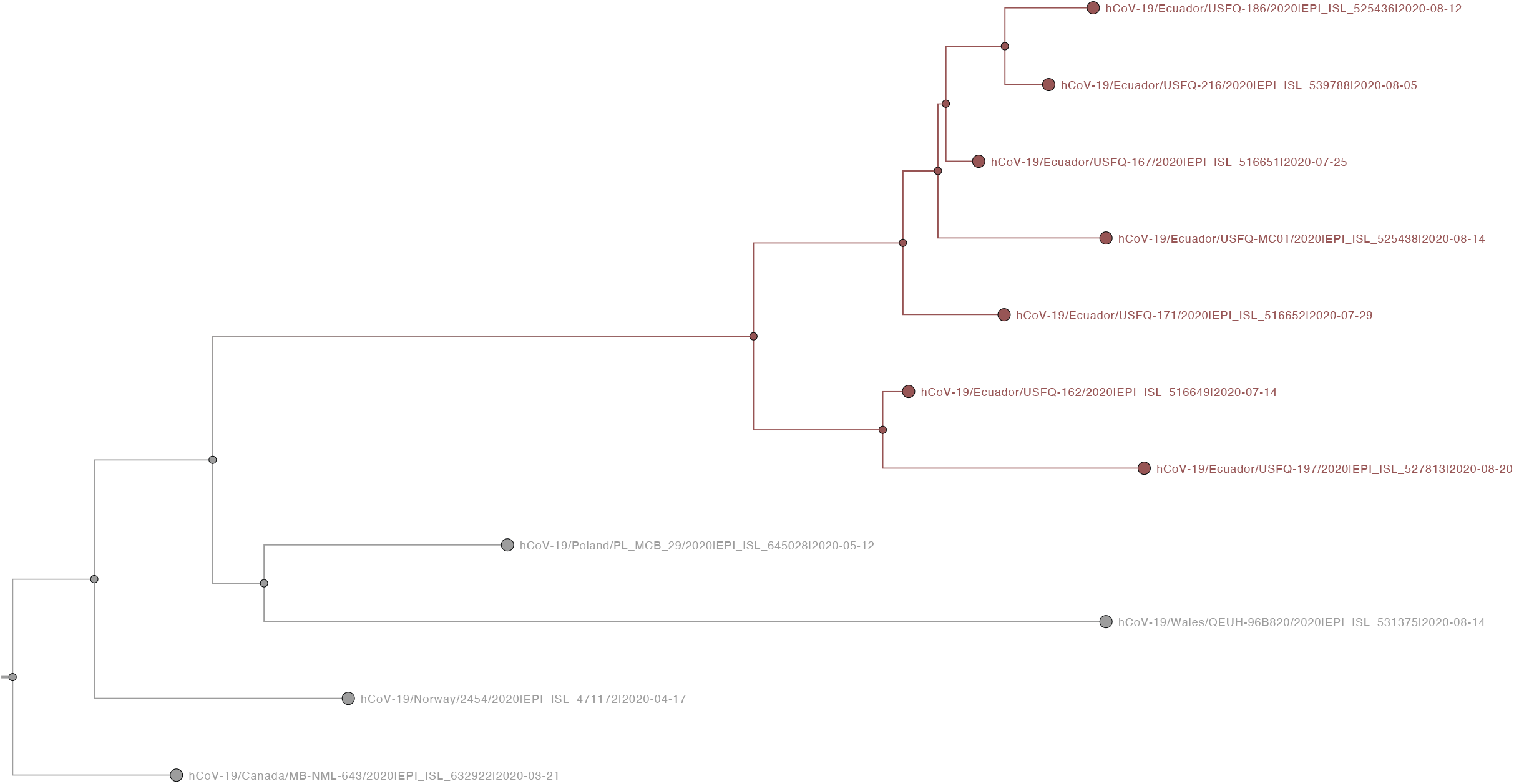
Subtree from the MCC tree, corresponding to transmission lineage O.

**Fig S7.**
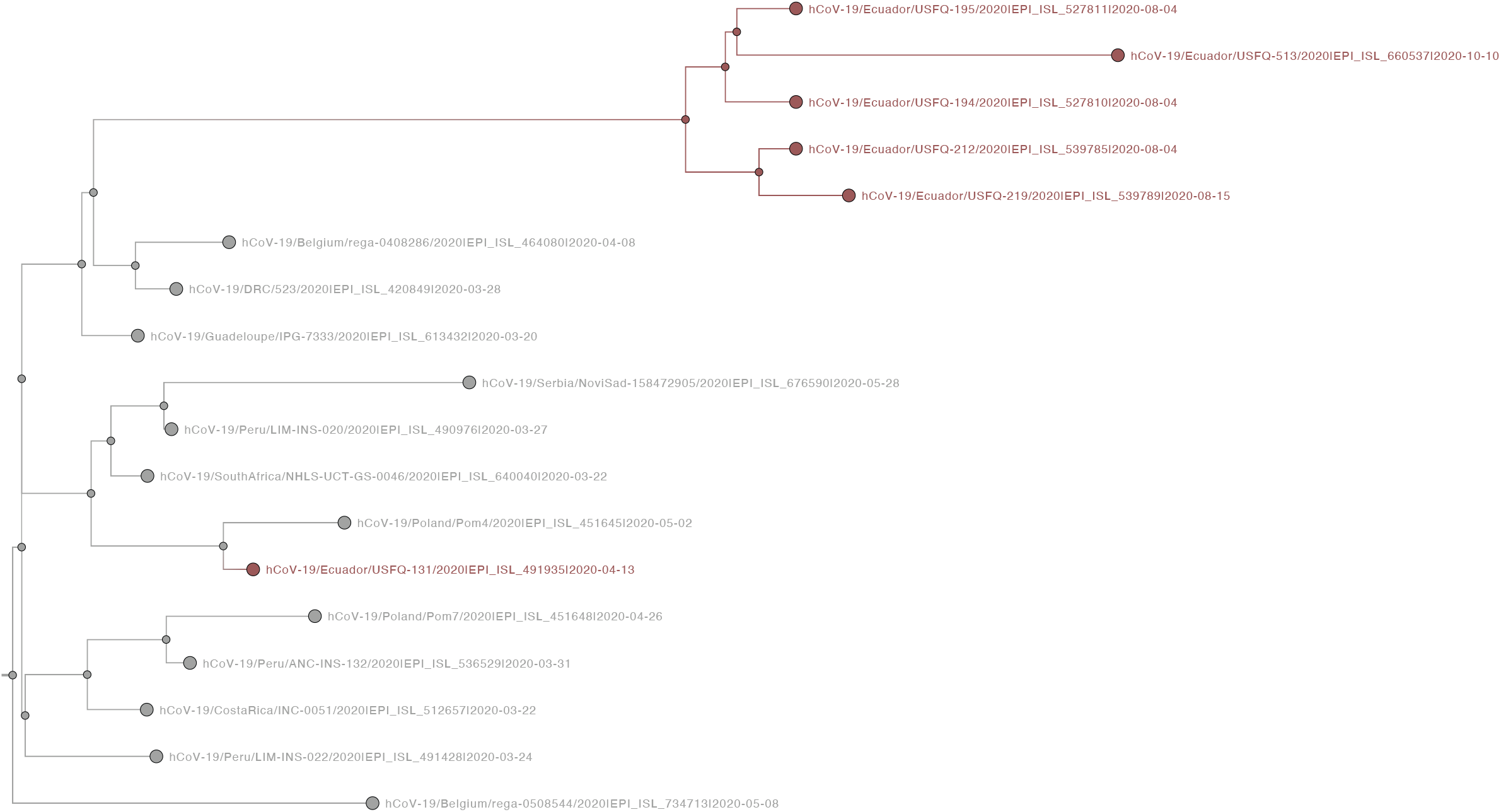
Subtree from the MCC tree, corresponding to transmission lineage F.

**Fig S8.**
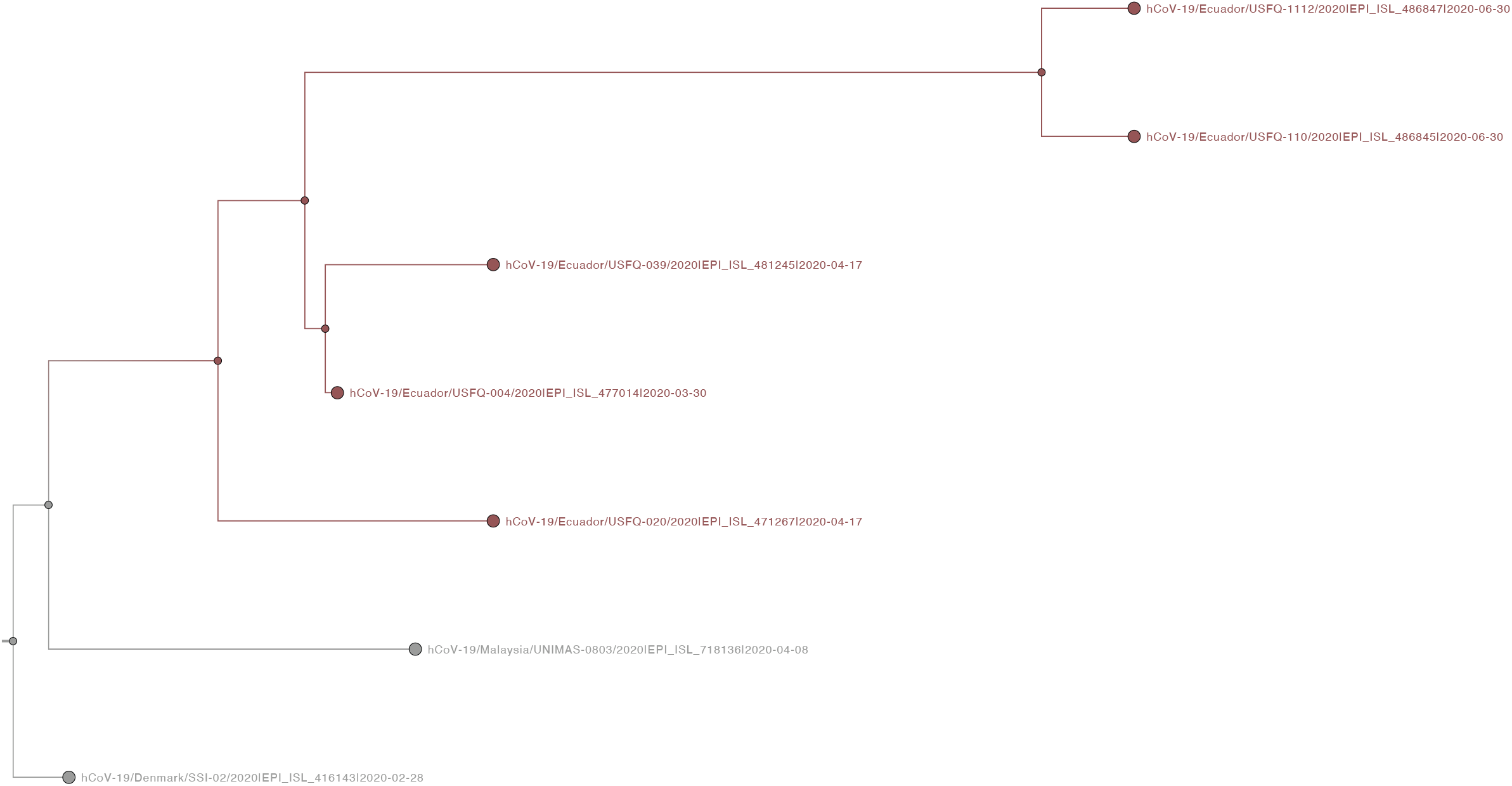
Subtree from the MCC tree, corresponding to transmission lineage B.

**Fig S9.**
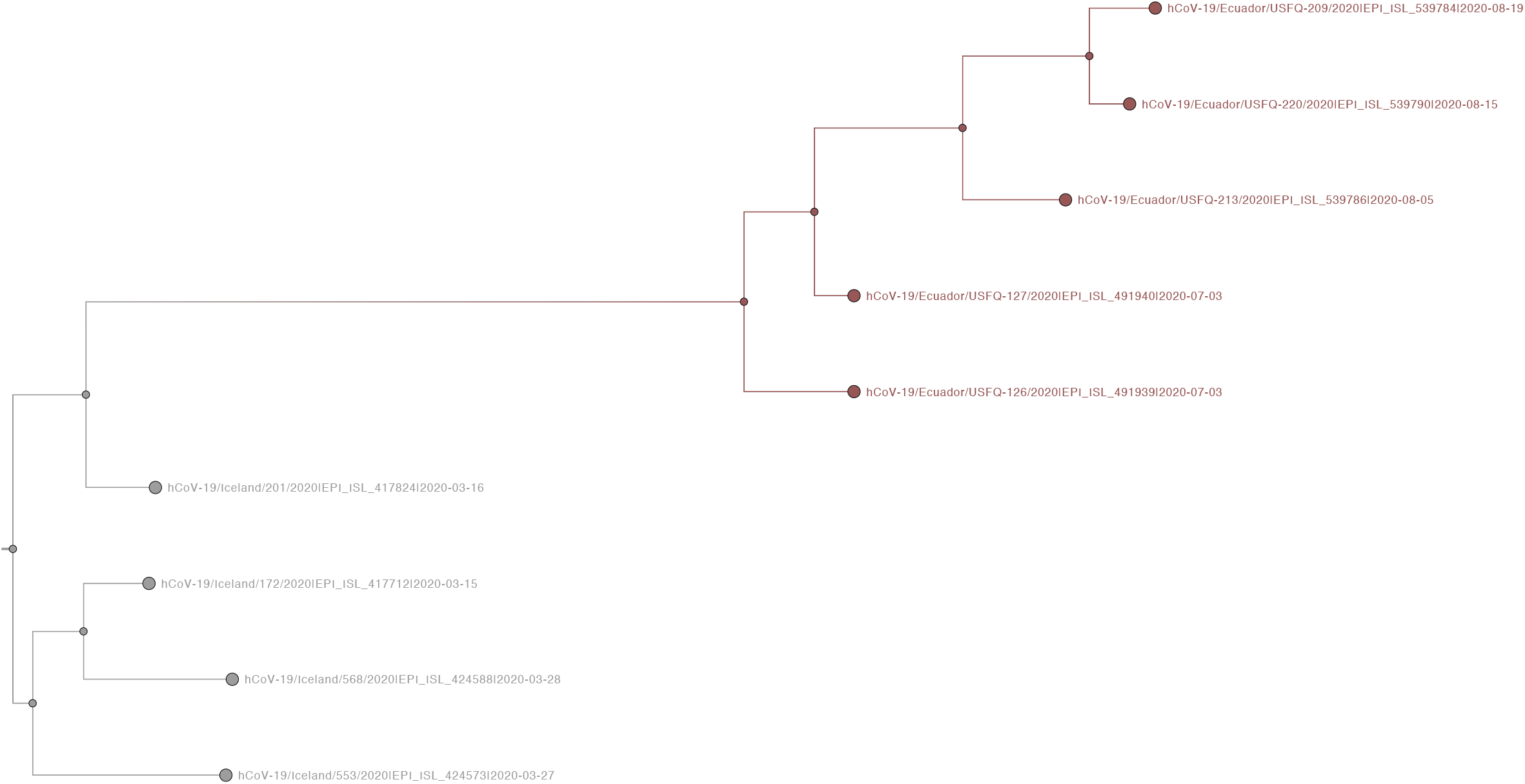
Subtree from the MCC tree, corresponding to transmission lineage M.

**Fig S10.**
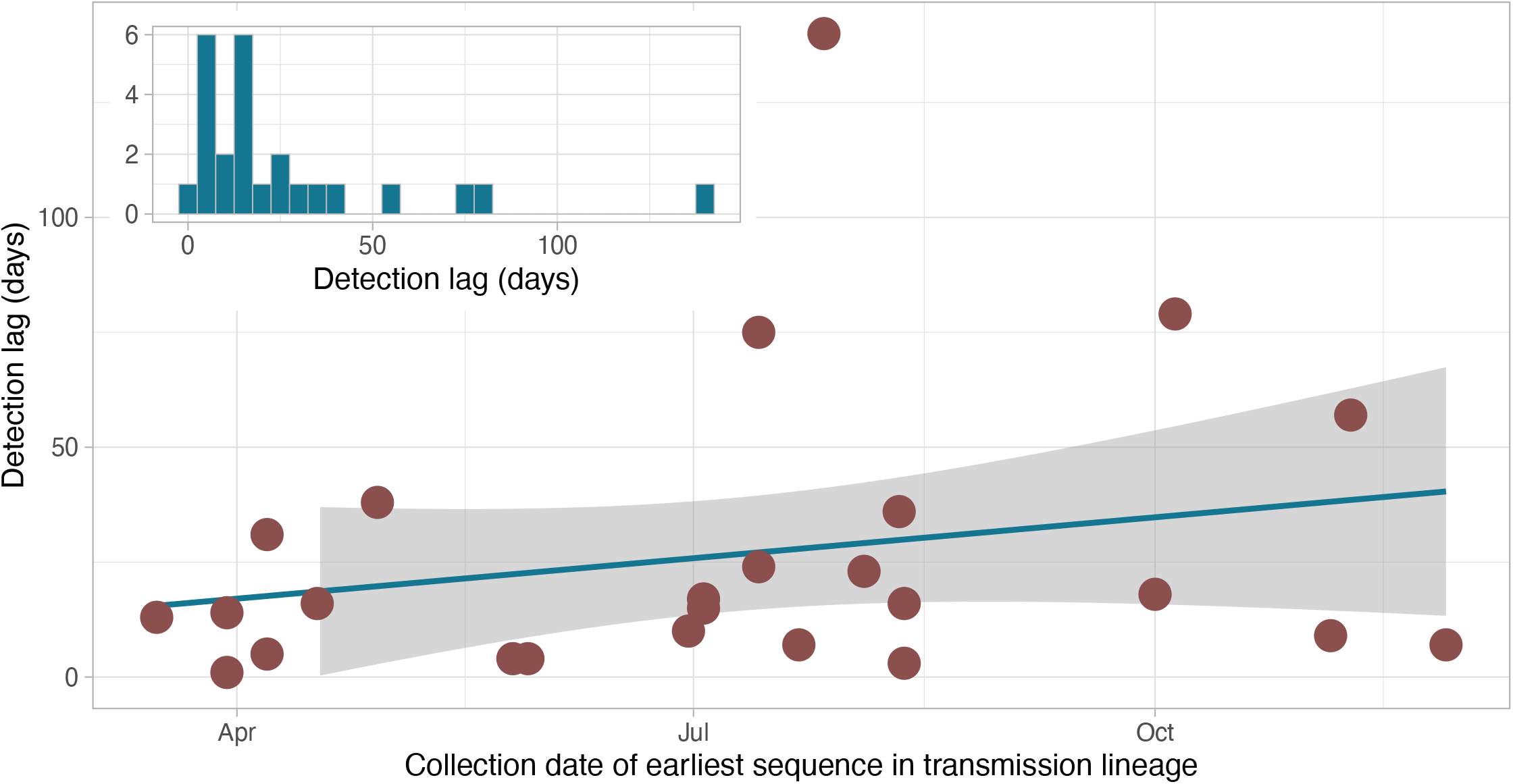
Changes in detection lag over time. Figure shows the detection lag of every transmission lineage (time between the TMRCA of a transmission lineage and the collection date of its earliest sequence), plotted against the collection date of the earliest sequence in said lineage. The inset shows the distribution of detection lag times in the data set.

**Fig S11.**
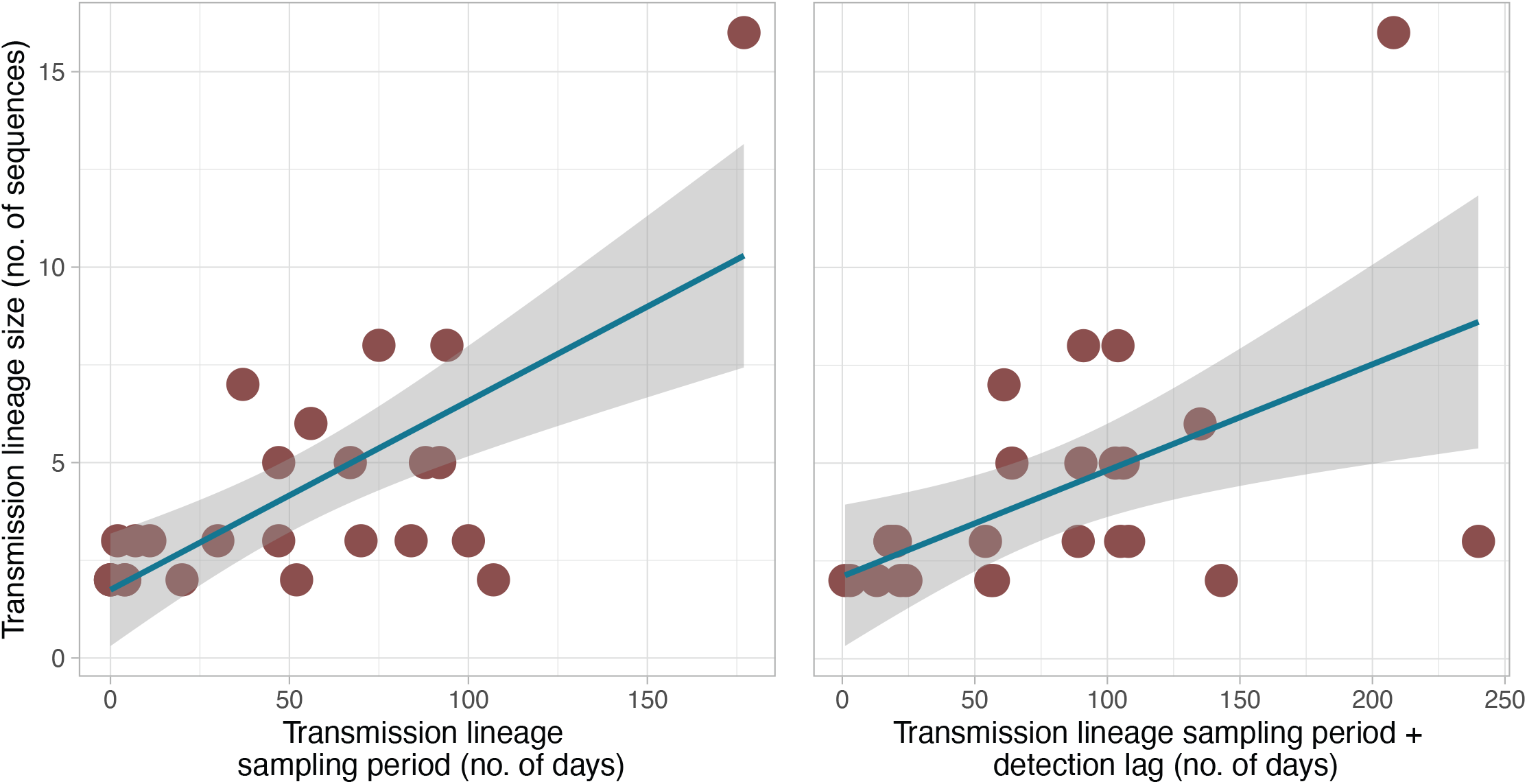
Comparison between the size of transmission lineages (i.e. number of sequences assigned to a transmission lineage) and their persistence in time estimated as either the number of days between the earliest and most recent sequences in said lineage (left) or the number of days between the transmission lineage TMRCA and the most recent sequence (right).

**Fig S12.**
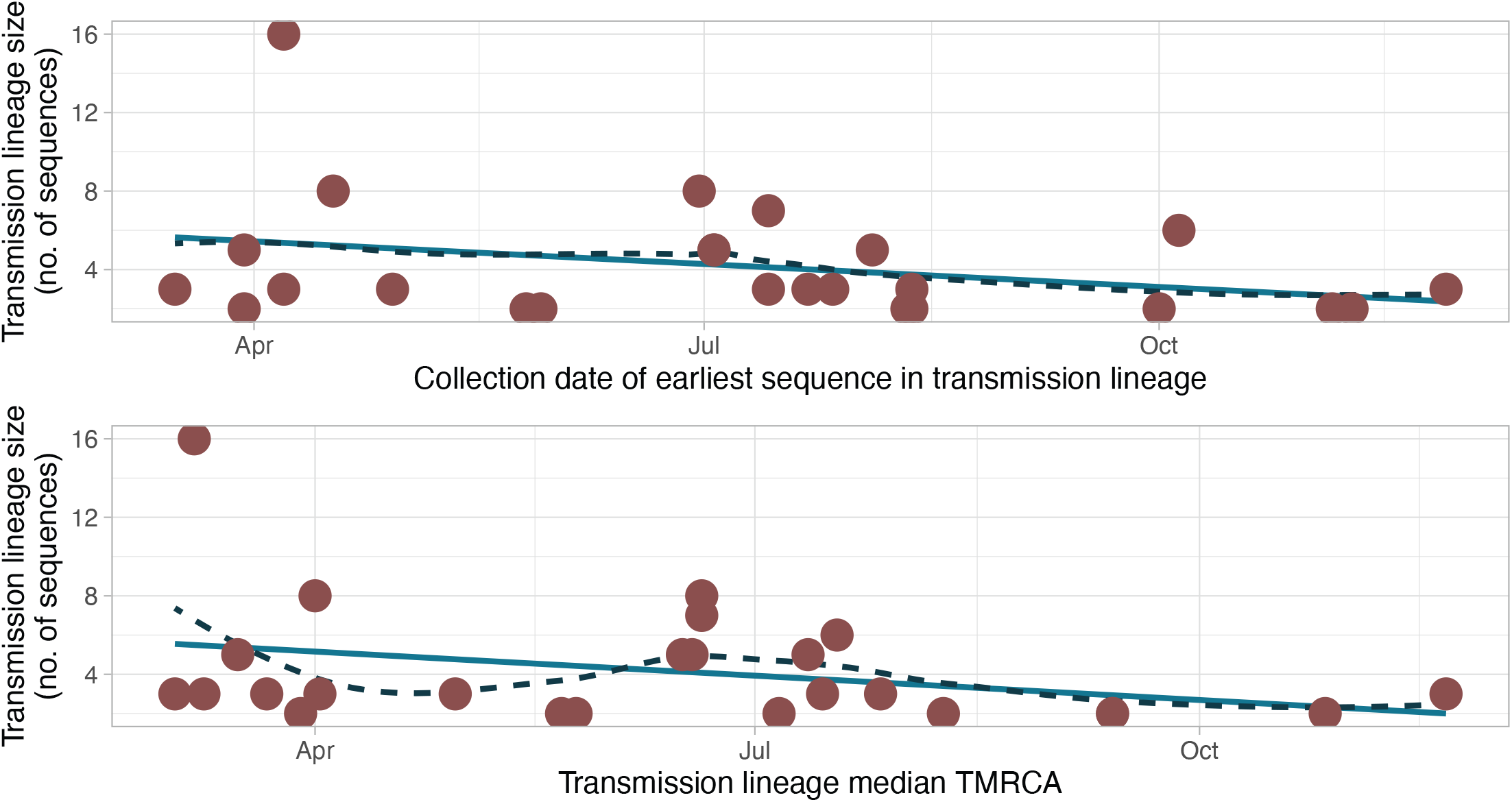
Comparison between the size of transmission lineages (i.e. number of sequences assigned to a transmission lineage) and the collection date of the first sequence identified in said lineage (upper panel) or the TMRCA for said transmission lineage (lower panel). Trend lines show a linear regression (green) and a fitted local polynomial regression (R function stats::loess, black and dashed).

**Fig S13.**
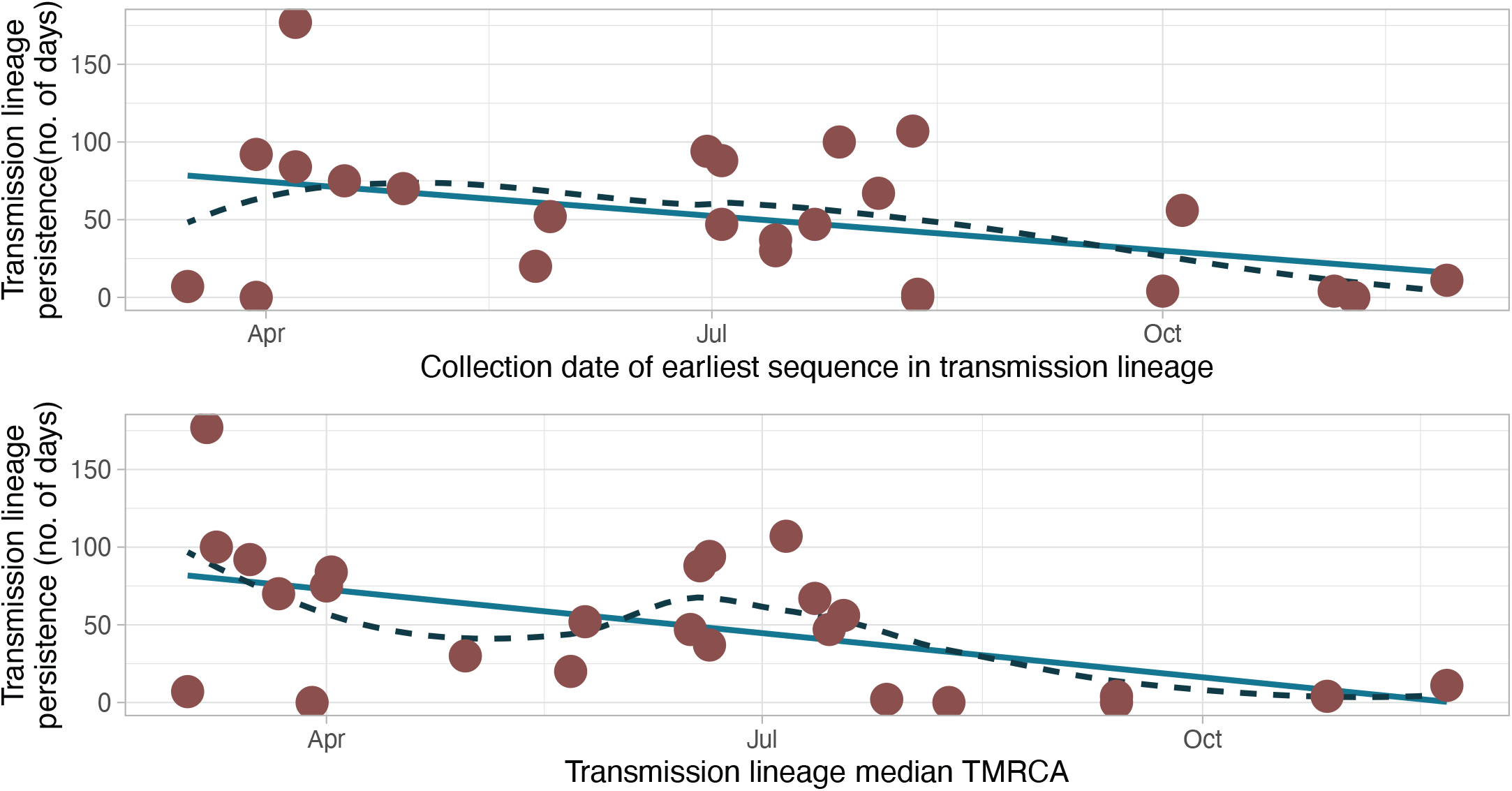
Comparison between the persistence in time of transmission lineages (number of days between the earliest and most recent sequences in said lineage) and the collection date of the first sequence identified in said lineage (upper panel) or the TMRCA for said transmission lineage (lower panel). Trend lines show a linear regression (green) and a fitted local polynomial regression (R function stat::loess, black and dashed).

**Fig S14.**
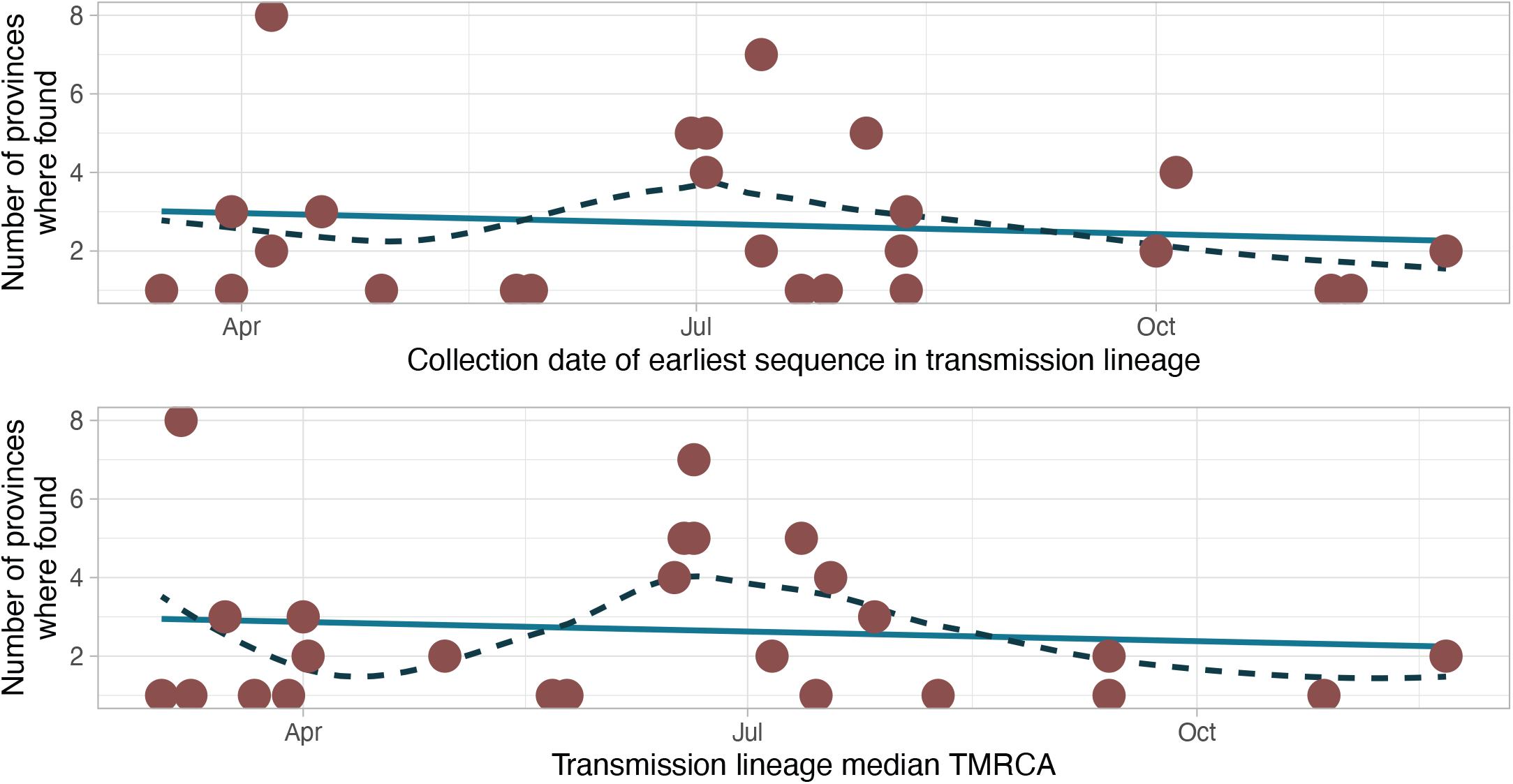
Comparison between the geographic spread of transmission lineages (number of provinces where they’ve been detected) and the collection date of the first sequence identified in said lineage (upper panel) or the TMRCA for said transmission lineage (lower panel). Trend lines show a linear regression (green) and a fitted local polynomial regression (R function stat::loess, black and dashed).

**Fig S15.**
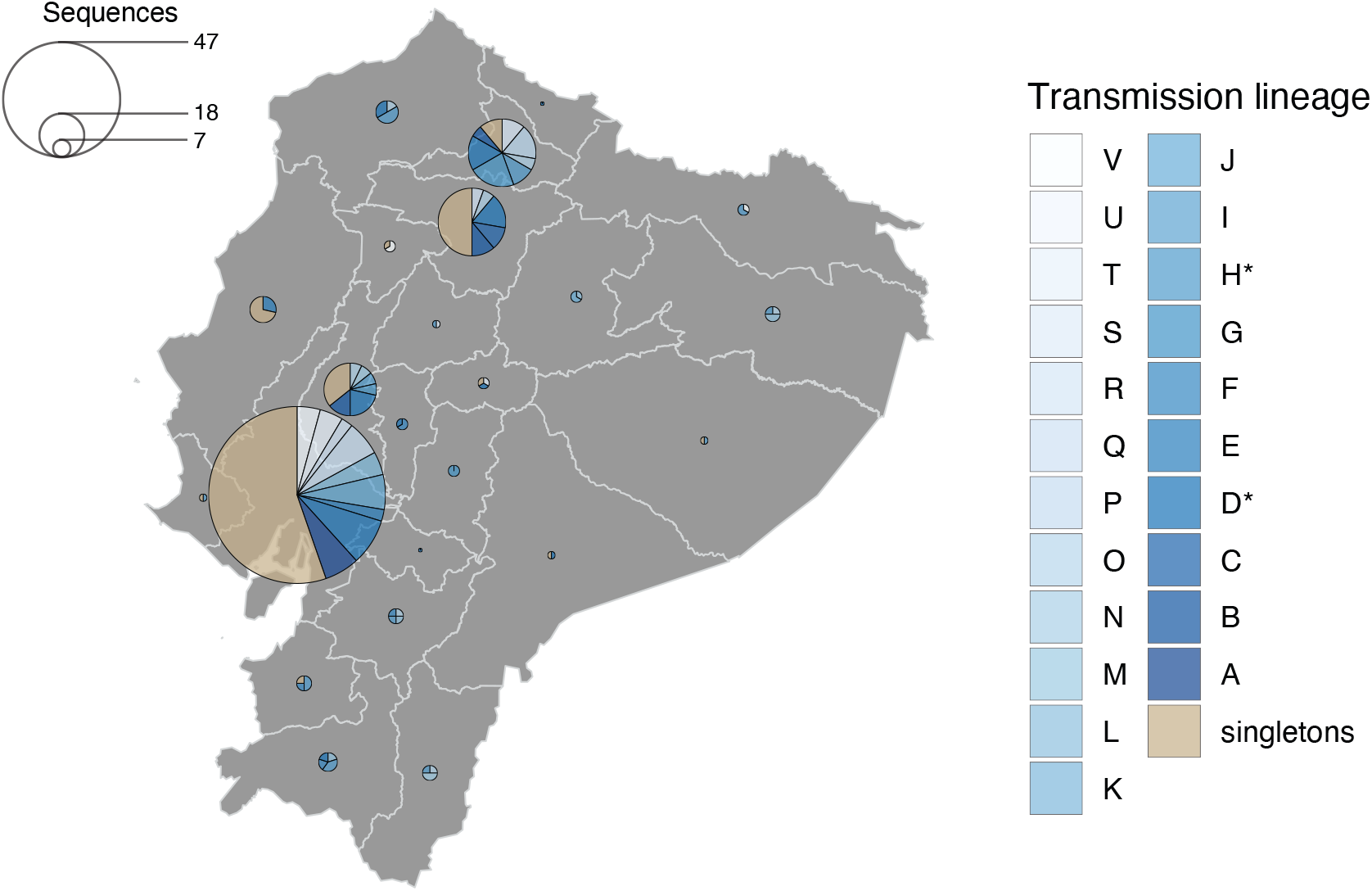
Map of Ecuador showing the contribution of individual transmission lineages (shades of blue) and sequences not associated with them (singletons) in each province. Circle radii represent the number of sequences per province.

**Fig S16.**
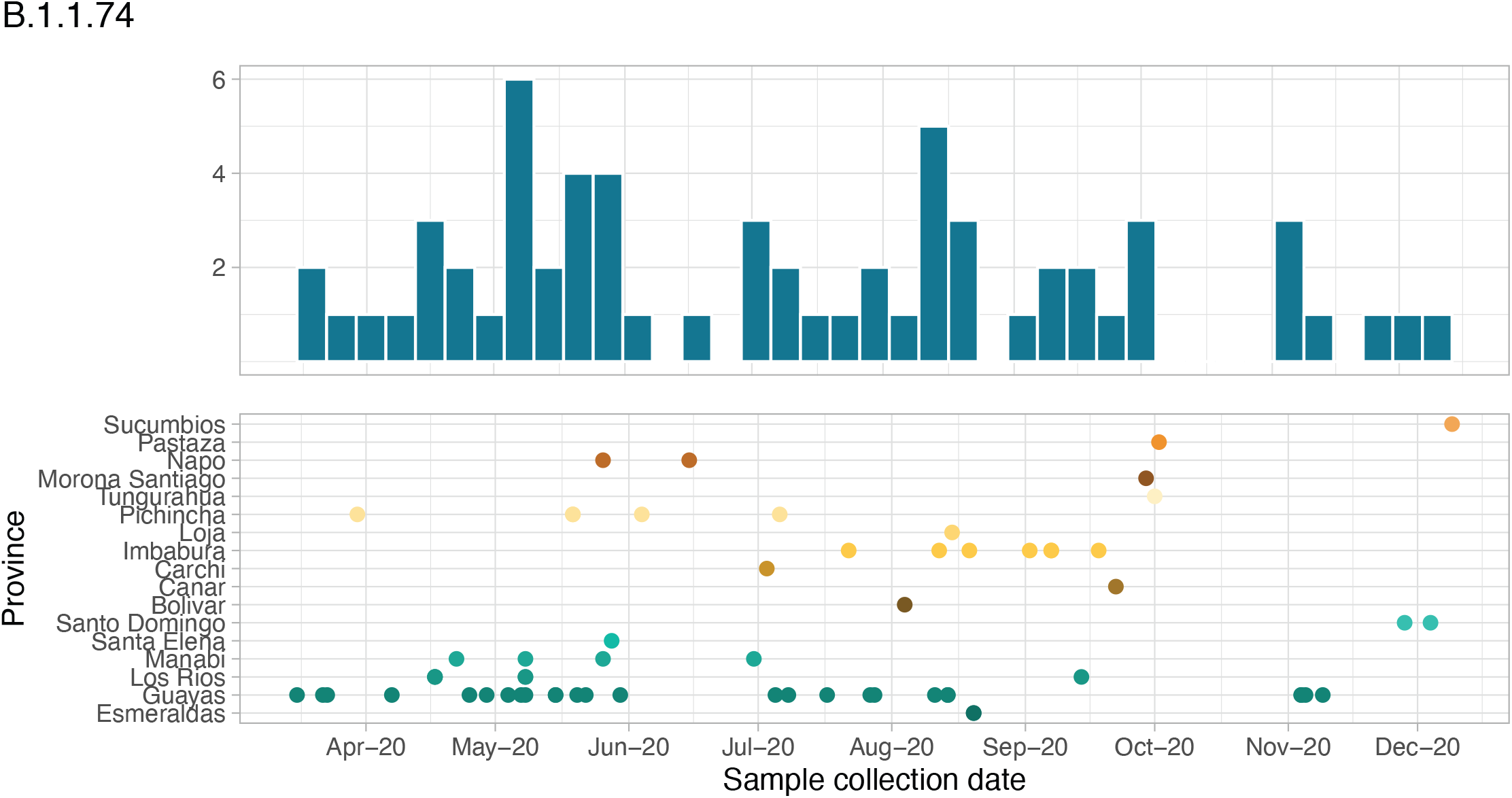
Summary of the identification of Pango lineage B.1.1.74 in Ecuador. The upper panel shows the number of sequences assigned to B.1.1.74 per 7-day time period, and the lower panel shows the province where these sequences were identified by sample collection date. Dot colours correspond to the province where the samples were collected.

